# Integration of Machine Learning to Identify Diagnostic Genes in Leukocytes for Acute Myocardial Infarction Patients

**DOI:** 10.1101/2023.09.07.23295181

**Authors:** Lin Zhang, Yue Liu, Kaiyue Wang, Xiangqin Ou, Jiashun Zhou, Houliang Zhang, Min Huang, Zhenfang Du, Sheng Qiang

## Abstract

**Background:** Acute myocardial infarction (AMI) has two clinical characteristics: high missed diagnosis and dysfunction of leukocytes. Transcriptional RNA on leukocytes is closely related to the course evolution of AMI patients. We hypothesized that transcriptional RNA in leukocytes might provide potential diagnostic value for AMI. Integration machine learning (IML) was first used to explore AMI discrimination genes. The following clinical study was performed to validate the results.

**Methods:** A total of four AMI microarrays (derived from the Gene Expression Omnibus) were included in this study (220 sample size), and the controls were identified as patients with stable coronary artery disease (SCAD). At a ratio of 5:2, GSE59867 was included in the training set, while GSE60993, GSE62646, and GSE48060 were included in the testing set. IML was explicitly proposed in this research, which is composed of six machine learning algorithms, including support vector machine (SVM), neural network (NN), random forest (RF), gradient boosting machine (GBM), decision trees (DT), and least absolute shrinkage and selection operator (LASSO). IML had two functions in this research: filtered optimized variables and predicted the categorized value. Furthermore, 40 individuals were recruited, and the results were verified.

**Results:** Thirty-nine differentially expressed genes (DEGs) were identified between controls and AMI individuals from the training sets. Among the thirty-nine DEGs, IML was used to process the predicted classification model and identify potential candidate genes with overall normalized weights >1. Finally, Two genes (AQP9 and SOCS3) show their diagnosis value with the area under the curve (AUC) > 0.9 in both the training and testing sets. The clinical study verified the significance of AQP9 and SOCS3. Notably, more stenotic coronary arteries or severe Killip classification indicated higher levels of these two genes, especially SOCS3. These two genes correlated with two immune cell types, monocytes and neutrophils.

**Conclusion:** AQP9 and SOCS3 in leukocytes may be conducive to identifying AMI patients with SCAD patients. AQP9 and SOCS3 are closely associated with monocytes and neutrophils, which might contribute to advancing AMI diagnosis and shed light on novel genetic markers. Multiple clinical characteristics, multicenter, and large-sample relevant trials are still needed to confirm its clinical value.

## 1 Introduction

Acute myocardial infarction (AMI), the most severe form of cardiovascular disease, is associated with [1, 2] millions of deaths annually around the world [3, 4]. Generally, the diagnosis of AMI includes clinical syndrome, electrocardiogram, and serum changes in enzyme levels [5]. However, AMI is easily misdiagnosed because of the following three aspects: nonclassic clinical symptoms [6, 7], atypical underappreciation [8], and an untimely serum peak. Because of the above three problems, a previous study [9] reported that the missed diagnosis rate of AMI is higher than 0.9%. The diagnosis and treatment of AMI must be prompt; otherwise, it may trigger irreversible results. Therefore, exploring new markers of AMI to decrease missed diagnoses is essential and urgent.

Leukocytes play an important and varied role in the entire evolution of AMI. During the acute injury phase of AMI, leukocytes promote a severe inflammatory cascade response through the polarization of M1 macrophages [10]. During the repair phase of AMI, M2 macrophages in leukocytes suppress inflammation and mediate the repair of injured myocardium [11]. Furthermore, leukocyte alteration positively correlates with AMI severity and, inversely, with patient survival [12, 13].

RNAs are involved in the evolution of AMI. For example, miR-155 correlated positively with the concentration of inflammatory cytokines - IL-6 and TNF-α [14] in AMI. Neutrophil-derived S100A8/A9 amplify granulopoiesis and cardiac injury in AMI mice [15]. Conversely, M2 macrophage-derived exosomes carry miR-1271-5p [16] to alleviate AMI-related cardiac injury. In conclusion, RNA on leukocytes plays a different role in the evolution of AMI, possibly related to different leukocyte subtypes. However, numerous studies have focused on integrating target interventions [12, 17] and leukocyte complications [17, 18]. Few studies have focused on the diagnostic value of leukocytes’ RNA. Because the leukocytes’ RNA is involved in the evolution of AMI, these RNA might have diagnosing value for AMI patients. The diagnosis value might be related to various leukocyte subtypes.

Machine learning (ML) helps humans learn patterns from complex data to predict future behavioural outcomes and trends. ML was widely utilized in variable filtering. A previous study used a single ML algorithm or two integrated ML algorithms (e.g., support vector machine [18] or least absolute shrinkage and selection operator [19]) to optimize variables. Still, these approaches may have missed potential genes [20]. Compared with a single ML algorithm, the integrated ML (IML) approach [21–23] we developed is more advantageous in variable screening and model building. IML helps identify potential genes mistakenly deleted by a single ML and find more meaningful variables [21]. IML integrates the advantages of a single ML, and its predictive classification value is better [23]. Based on a favourable filtration value in transcriptomics of IML, IML might be used to comprehensively explore the diagnostic value in AMI patients.

In summary, we aim to explore the potential diagnostic value of transcriptome within leukocytes for identifying AMI patients. Because of IML’s good variable screening and excellent predictive value, IML was first used to mine diagnostic genes in AMI leukocytes with multiple microarrays. Single microarray data might have inherent biases in capturing the entire transcriptomic landscape, so multiple microarrays are integrated after resolving batch effects to reduce bias and validate each other. And clinical validation was added to confirm the result. The relationship between transcriptome and leukocyte subtypes was unclear, so the correlation between immune cells and target transcriptome was subsequently accomplished. We expect to explore the functional roles of the identified genes in AMI pathophysiology, investigating their potential as therapeutic targets.

## 2 Methods

### 2.1 Data acquisition

The raw data were obtained from the Gene Expression Omnibus (GEO, March 27, 2022). AMI patients have similar symptoms to SCAD patients, which were set as the controls. An increasing leukocyte may influence the result of other cardiovascular diseases (*e.g*., stroke [24, 25] and heart failure [26]), to be excluded. AMI is easily misdiagnosed as SCAD. Leukocytes are also altered in other cardiovascular diseases. Based on the above, the following inclusion and exclusion criteria were set: I) inclusion criteria—(i) diagnosed as AMI patients on admission; (ii) transcriptome was obtained from leukocytes in blood; (iii) initial data were free and accessible; and (iv) the control individuals were diagnosed with stable coronary artery disease (SCAD); and II) exclusion criteria—(i) other cardiovascular diseases suspected and (ii) blood were taken more than one day after hospitalization.

### 2.2 Data processing

To ensure the reliability of the data, the R package *sva* (version 3.46.0) was applied to data integration to minimize the branch effects with the *ComBat* function and parametric adjustments. Regarding the distribution ratio of previous literature (1.64:1 [27] to 5:1 [28]) and to minimize the branching effect, this research was distributed in the training or testing sets at a ratio of 5:2. GSE59867 was included in the training set. In contrast, GSE60993, GSE62646, and GSE48060 were included in the testing set. In brief, the training set was applied to explore candidate diagnostic genes, and the testing set was used for validation. Based on the differential DEGs, three functional enrichment analyses were developed via the Kyoto Encyclopedia of Genes and Genomes Gene Set Enrichment Analysis (KEGG-GSEA), Gene Ontology (GO), and Disease Ontology (DO). In addition, the GO terms included three branches: molecular function (MF), biological process (BP), and cellular components (CC). Notably, the novel IML served two functions: developing classification ML and exploring the candidate variable. Finally, the above candidate genes were verified in the testing group and clinical study, and an immune analysis among the candidate genes was performed. CIBERSORT was processed for immune correlation analysis in the *corrplot* R package (version 0.92). And the primary code was link with https://github.com/Linzhang-BiuBiuBiu/ML-for-diagnosis-genes..git.

### 2.3 Searching for DEGs

Because the same gene may have multiple sequences, the transcriptome will appear to have several expression data for the same genes. For the same genes, *limma* (version 3.54.0) was employed to identify the DEGs with the average gene expression. According to the Benjamini and Hochberg method, two thresholds were established: an absolute value of fold change (|logFC|) >0.7 (previous studies were 0.5 [29]-1 [23]) and a false discovery rate [30] <0.05.

### 2.4 IML of six ML algorithms

Classification models of IML, composed of six ML algorithms, were processed, covering support vector machine (SVM), neural network (NN), random forest (RF), gradient boosting machine (GBM), decision trees (DT), and least absolute shrinkage and selection operator (LASSO). In brief, IML was used to identify candidate genes with the overall normalized weights. The six ML algorithms were developed to optimize parameter settings, model development in the training sets, and validation in the testing sets. For stability, all ML algorithms were tenfold cross-validated. Notably, an accuracy value was applied to evaluate the predictive classification value, and a higher accuracy value showed a better classification value of the six ML algorithms.

LASSO [31] minimizes the sum of squares of the residuals when the sum of the absolute values of the regression coefficients is less than a constant, producing specific regression coefficients equal to 0 and filtering variables. LASSO was processed with the *glmnet* (version 4.1-6) R package. *cv.glmnet* was utilized to majorize lambda. For the parameters, the scale of “lambda” was set between 0 and 100 with “binomial” and “class”.Based on the minimum lambda, *glmnet* was processed to the LASSO with alpha and a “binomial” method in training sets.

SVM aim to find the separating hyperplane [32] that divides the dataset correctly with the largest geometric interval. SVM was developed with the *e1071* R package (version 1.7–12). *tune.svm* was adopted to optimize the settings parameter with the kernel of “linear”, and the cost between 1 and 20.

DT [33] is based on a tree structure that judges (one or more) sample attributes sequentially, from top to bottom, up to the leaf nodes of the decision tree and derives the final result. DT was processed with *rpart* (version 4.1.19) and *rpart.plot* (version 3.1.1). Based on the “class” method and a cp value of 0.001, the *rpart* function was adopted for the DT model.

RF use a “ bagging “ technique [34] to construct complete decision trees in parallel by randomly self-sampling dataset samples and features. RF was completed with the R package *randomForest* (version 4.7-1.1). First, the *tuneRF* function was adopted to optimize 0-700 trees with one step size. RF was developed based on the minimum error rate to optimize the number of trees.

NN outputs model [35] by inputting multiple nonlinear models and weighted interconnections between various models. NN was processed with *neuralnet* (version 1.44.2) with *neuralnet* function, five layers (an input, an output, and three hidden layers), err.fct of “sse”, and the linear.

GBM serially generates a series of weak learners [36], which are directly used to form the final model by combining them. Compared with the other 5 ML algorithms, GBM processed more steps and was prone to making mistakes. The GMB was developed with *h2o* (version 3.38.0.1).

First, the Java operating environment was installed, which is the virtual environment of GBM. Essential for running the memory setting in *h2o.init*, the model memory of GBM was adjusted to 8G. The h2o data type in GBM was inevitable, and the *as.h2o* function was utilized to transform the data format. Next, *h2o.gbm* tuned the parameters and developed the model with the “Bernoulli” distribution, 200 trees, a learning rate of 0.001, and a sample rate of 90%.

Furthermore, with the weights of the above six ML algorithms in DEGs, the normalized sum weight of IML was calculated as follows: overall weights = abs(RF)/abs(RFmax) + abs(SVM)/abs(SVMmax) + abs(LASSO)/abs(LASSOmax) + abs(NN)/abs(NNmax) + abs(GBM)/abs(GBMmax) + abs(DT)/abs(DTmax). For instance, if the weight of interleukin-6 in six ML algorithms was 30, −22, 20, −2, 320, and −8, the maximum absolute value weights in the six ML algorithms were 60, 88, 80, 8, 640, and 16. Therefore, the overall weight of interleukin-6 was |30|/60+|-22|/88+|20|/80+ |-2|/8+|320|/640+|-8|/16=2.25. With normalized overall weights >1, the candidate genes were estimated by the area under the curve (AUC).

### 2.5 Clinical validation

The clinical trial was performed according to the Declaration of Helsinki guidelines. All AMI and SCAD patients provided individual written informed consent from October 10 2022, to December 31 2022, and the Ethics Review Committee of Jinghai District Hospital approved the study. There was no increase in the cost of treatment for the patients, no addition of other intervention in the treatment, and the blood samples used were taken from the discarded blood of the patients after their routine blood tests on the same day. If the patient did not have a routine blood test on that day, then the patient was excluded. All AMI patients underwent coronary angiography, and blood samples were collected in anticoagulant tubes on admission. Density gradient centrifugation [37, 38] was performed for leukocyte isolation. In brief, 8 mL of Ficoll solution was added to 8 mL of anticoagulated blood, and the upper plasma layer was discarded after centrifugation. The white cell layer at the isolate’s junction was aspirated, added to 10mL of saline, and centrifuged; the bottom layer was the leukocytes. RNA, isolated from leukocytes, was synthesized with reverse transcription kits (Takara, Shiga, Japan). Quantitative reverse transcription PCR was executed on an ABI7900HI (Thermo Fisher Scientific). According to previous literature, the relative content of the candidate genes was scaled to the reference gene (GAPDH [39]), and **Table 1** lists the primer sequences.

**Table 1.**
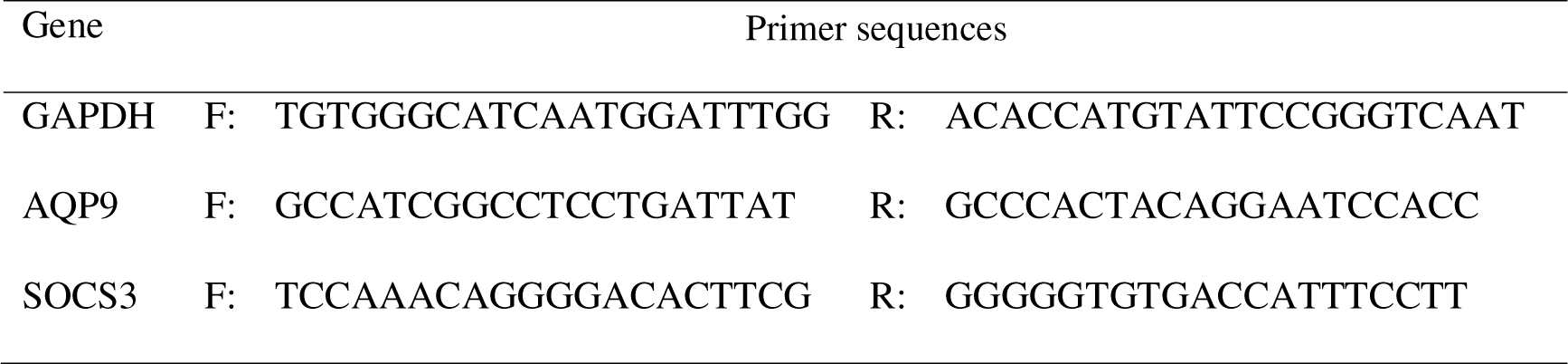
List of primers for real-time PCR analysis in GAPDH, AQP9, and SOCS3.

## 3 Results

### 3.1 Included datasets

A total of 4 datasets (**Table 2**) (220 sample sizes), namely, GSE59867, GSE60993, GSE62646, and GSE48060, were integrated for this study. The training set was obtained from GSE59867 (46 controls and 111 AMI patients) based on a raw ratio of 5:2. Furthermore, the testing set was integrated with the other three datasets (28 controls and 35 AMI patients), namely, GSE60993, GSE62646, and GSE48060. The following analysis is presented in **Fig 1**.

**Table 2.**
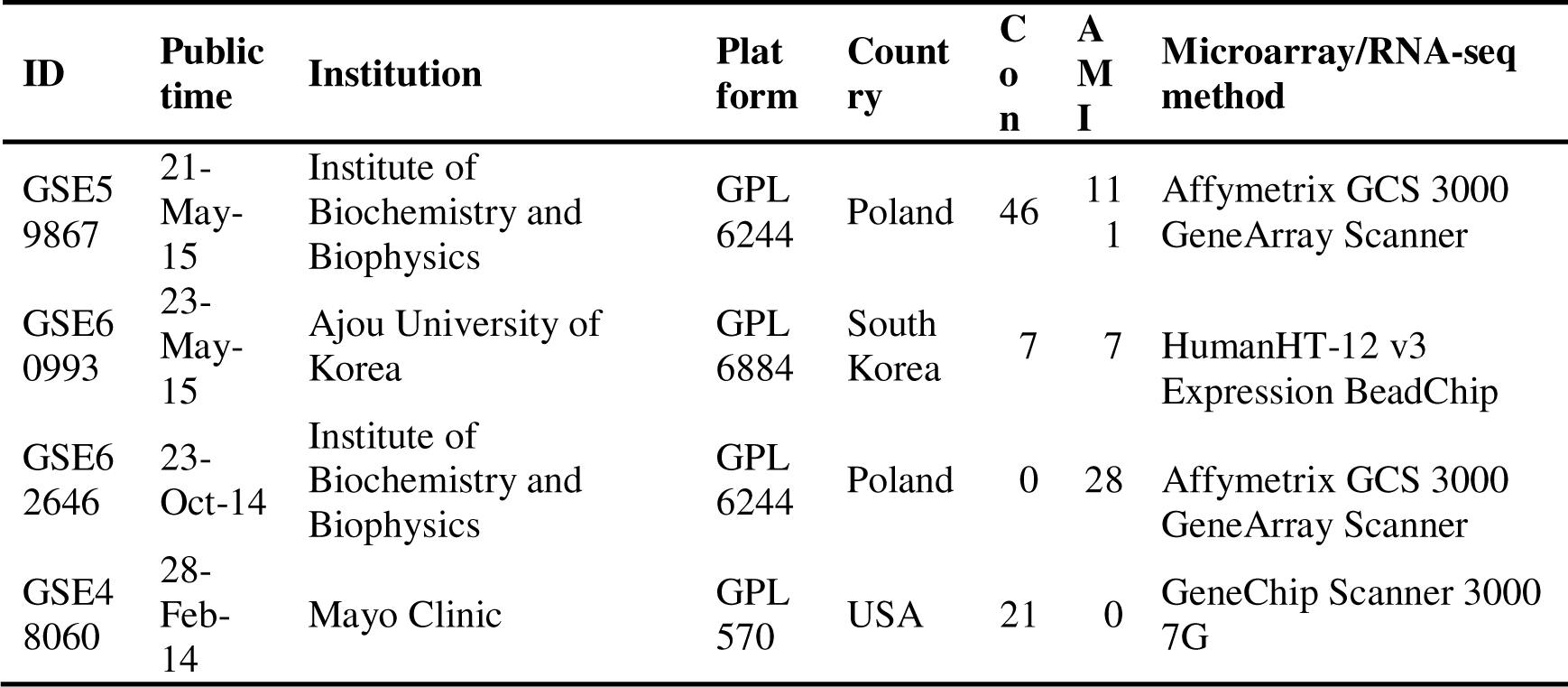
Fundamental information in the four datasets.

**Fig. 1.**
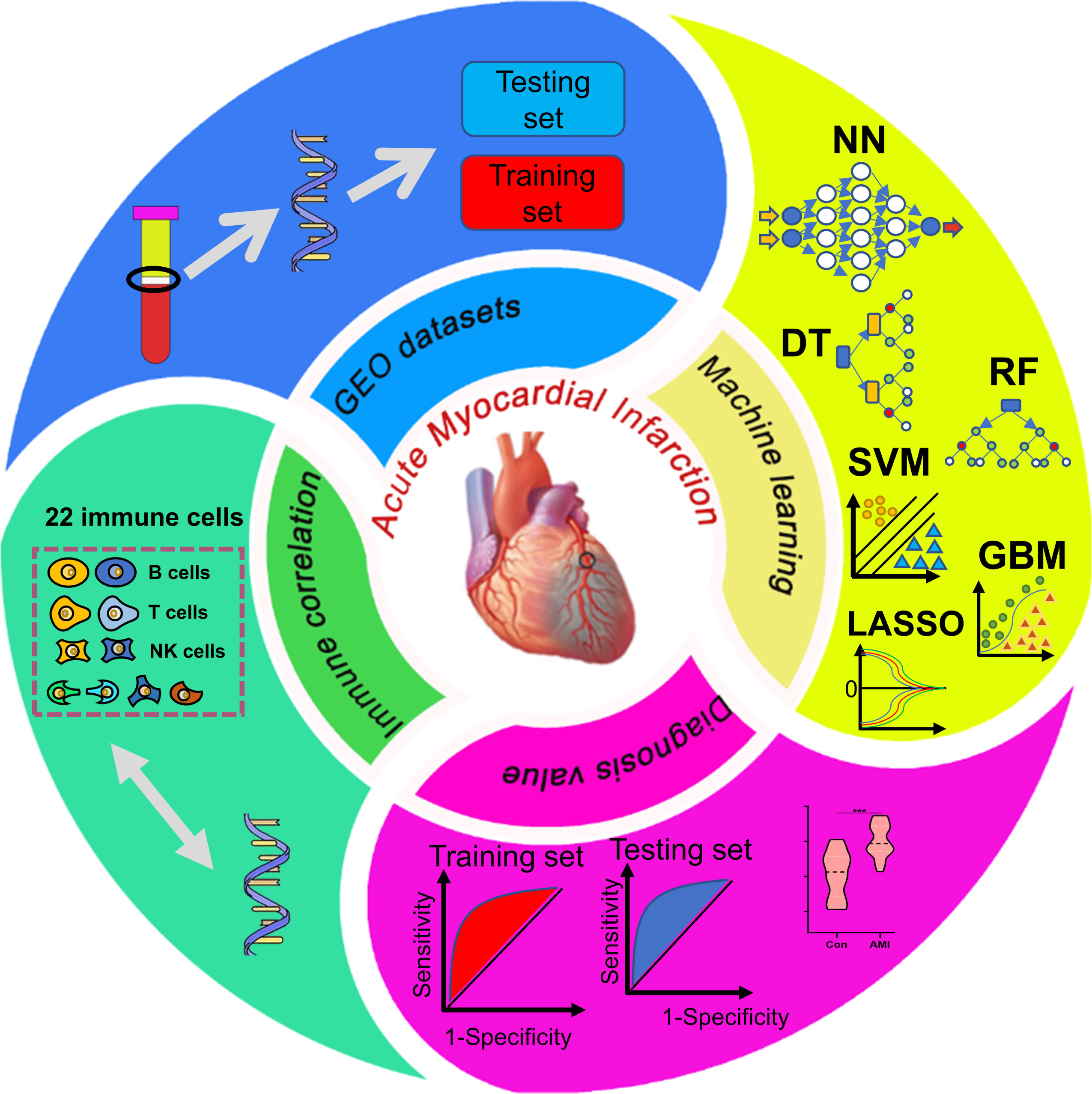
The workflow of this study contains four parts: GEO datasets for training and testing sets, machine learnings for classification and variable filtration, diagnosis value verification, and immune correlation.

### 3.2 DEG identification

Thirty-nine DEGs were identified (**Table S1**) in a training set from 17,049 RNAs. Compared to the control group (SCAD), 28 genes were upregulated (SOCS3, HP, ECRP, AQP9, FAM20A, CES1, STAB1, NRG1.1, NRG1, DYSF, RNASE1, RNASE2, ASGR2, CYP1B1, MERTK, FCGR1A.2, MIR21, FCGR1A.1, TCN2, VSIG4, PPARG, FCGR1A, SLED1, S100A9, FMN1.1, CD163, TMEM176A, and SERPINB2) and 11 genes were downregulated (KLRC3, KLRD1, KLRA1P, DTHD1, KLRC4, MYBL1, CLC, KLRC2, KLRC4-KLRK1, SNORD20, and SNORD45B) in AMI individuals (**Fig. 2**).

**Fig. 2.**
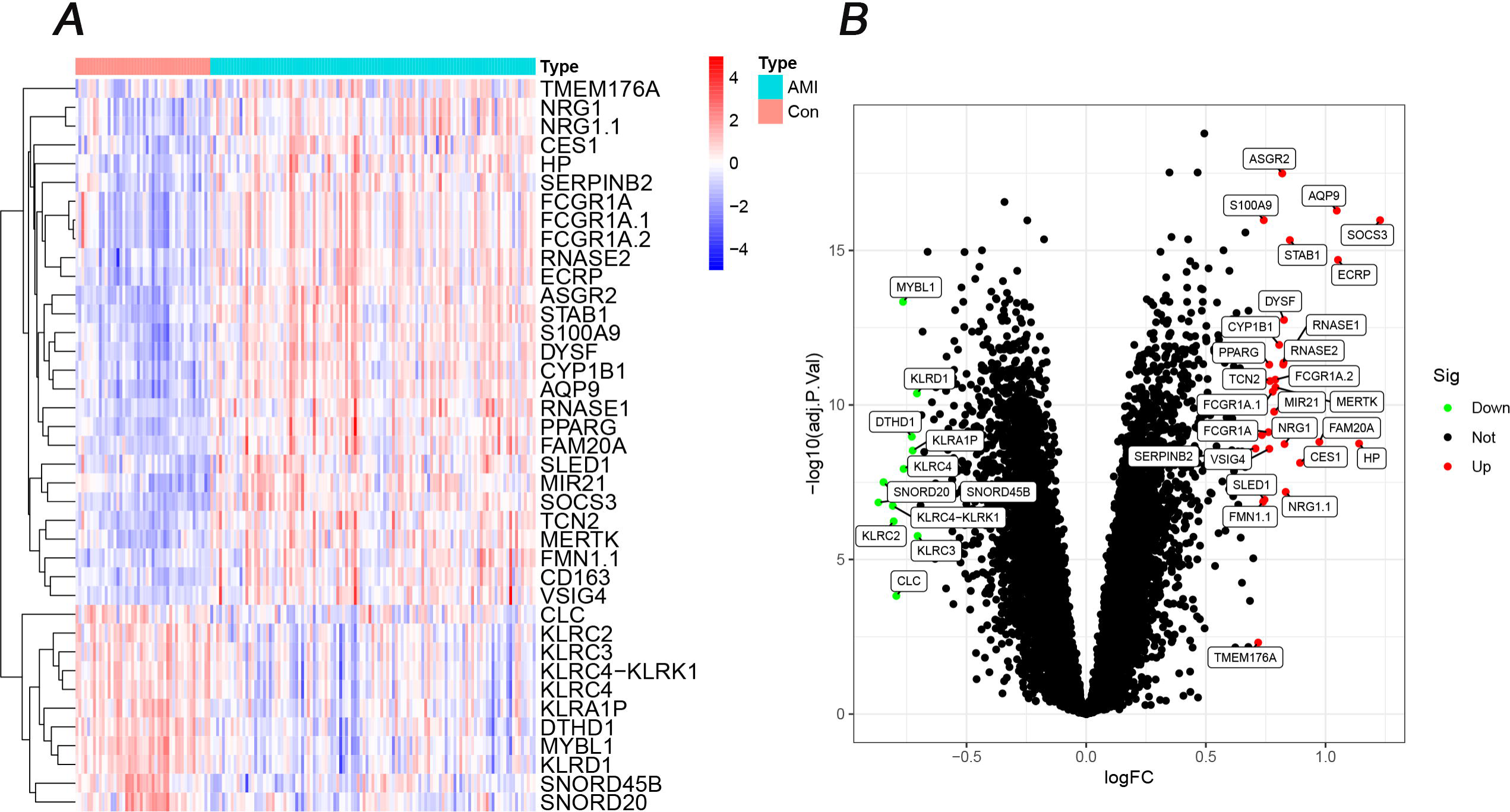
Heatmap and volcano plot of 39 DEGs in the AMI and control groups. **A** Red in the heat map indicates high expression, and a blue indicates low expression. **B** Green in the volcano map suggests lower expression, and red indicates high expression.

### 3.3 Functional analysis

Based on the above DEGs, 45 GSEA terms (**Table S2**) were identified, and the top 5 are shown in **Fig. 3A-B**; 160 GO terms (**Table S3**) were identified, and the top 5 are shown in **Fig. 3C**; and the top 10 of 57 DO terms (**Table S4**) are shown in **Fig. 3D**. In GSEA-KEGG of AMI, the top 3 were Fc gamma R-mediated phagocytosis, Huntington disease, and Leishmania infection. In GO, the top 3 in BP were the stimulatory C-type lectin receptor signalling pathway, response to lectin, and cellular response to lectin. In DO terms, the top 3 were atherosclerosis, arteriosclerotic cardiovascular disease, and arteriosclerosis.

**Fig. 3.**
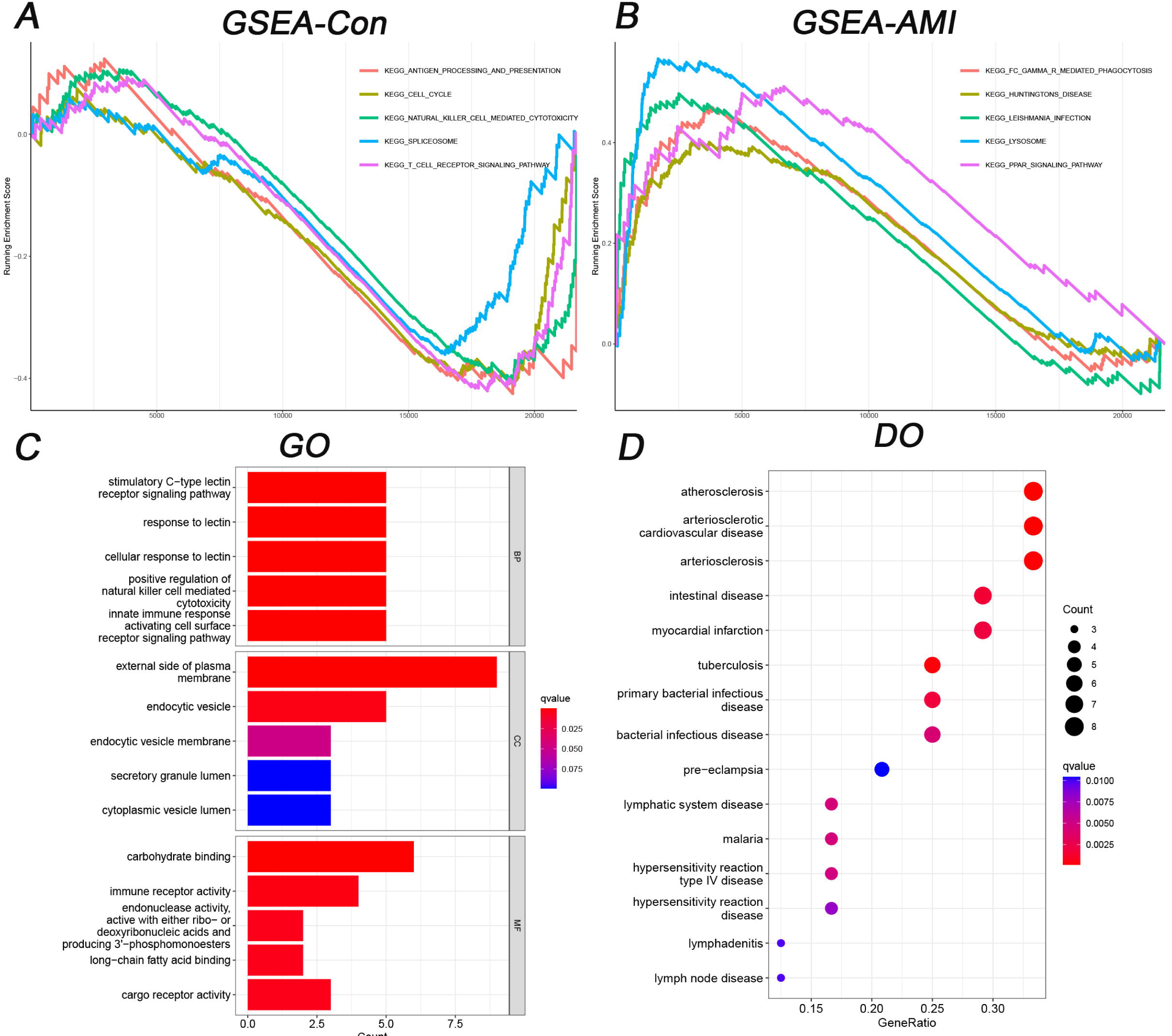
Functional analysis of GSEA, GO, and DO terms. **A** The top 5 GSEA-KEGG pathways in controls. **B** The top 5 GSEA-KEGG pathways in AMI patients. **C** The top 5 GO terms in BP, CC, and MF. **D** The top 15 DO terms.

### 3.4 IML of six ML algorithms

Six ML algorithms (**Fig. 4**) and their accuracies (**Table 3**) were assessed. Eight genes were identified in LASSO (**Fig. 4A**), and the training and testing sets’ accuracy value was 70.70% (**Table 3**). In SVM, 13 genes were filtered (**Fig. 4B**), and the accuracies were 88.46% and 91.84%, respectively. The error rate of RF (**Fig. 4C**) decreased with an increasing number of trees. Until 161 trees, the error rate of RF was minimized, and the accuracy of the two sets was 98.09% and 100%. In DT (**Fig. 4D**), the gene expression of 9.8 in AQP9 could discriminate the control and AMI groups, while the accuracies were unstable, 94.27%, and 75.52%. In GBM (**Fig. 4E**), 6-fold methods were established to optimize the diagnosis genes, but unstable accuracies, such as the above ML algorithms, were 93.30% and 85.71%. In the NN (**Fig. 4F**), although sufficient for discriminating the controls and AMI patients with three hidden layers, the accuracy was either 83.74% or 71.43%. Among the above ML algorithms, the raw weights of 39 DEGs were identified (**Table S5**). Interestingly, RF had the highest and most stable accuracy value among all ML algorithms. The normalized overall weights (**Table 4**) were calculated to filter the candidate variables. Twenty-six genes (ASGR2, SOCS3, AQP9, PPARG, RNASE1, DYSF, S100A9, FCGR1A, VSIG4, STAB1, MYBL1, KLRD1, ECRP, TCN2, FAM20A, MERTK, HP, RNASE2, DTHD1, CLC, SNORD20, CD163, NRG1, SNORD45B, CYP1B1, and KLRC2) were identified because of overall weights >1 (**Table 4**).

**Fig.4.**
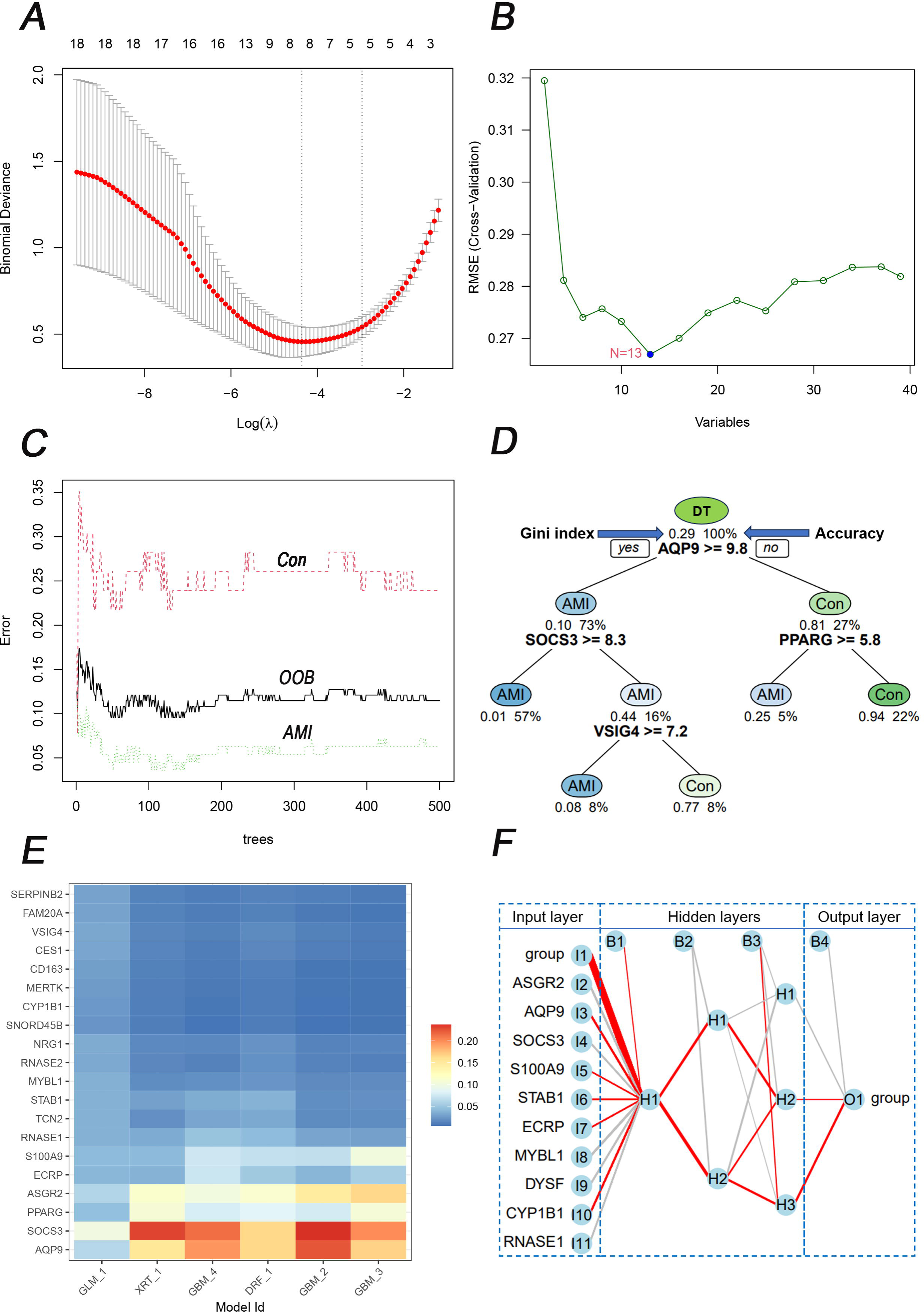
Six ML algorithms for classification with 39 DEGs. A LASSO for eight candidate genes and the error bars mean the fluctuation range of Binomial Deviance; B SVM for 13 candidate genes. C RF discriminated between the control and AMI groups. And The red, black, and green lines represent the Con, out-of-bag (OOB), and AMI groups respectively. D DT discriminated between the control and AMI groups. E A 6-fold GBM submodel was constructed. The heat map illustrates the importance of genes in each respective submodel. The intensity of the color corresponds to the significance of the gene in the particular submodel. F NN discriminated between the control and AMI groups. All 39 DEGs were involved in modeling in NN, and there are ten because of space limitations. If an edge is colored red, it indicates a positive correlation, meaning that the current feature positively affects the classification result. Conversely, if the edge is gray, it implies a negative correlation. Furthermore, the thickness of the edge signifies the weight’s magnitude.

**Table 3.**
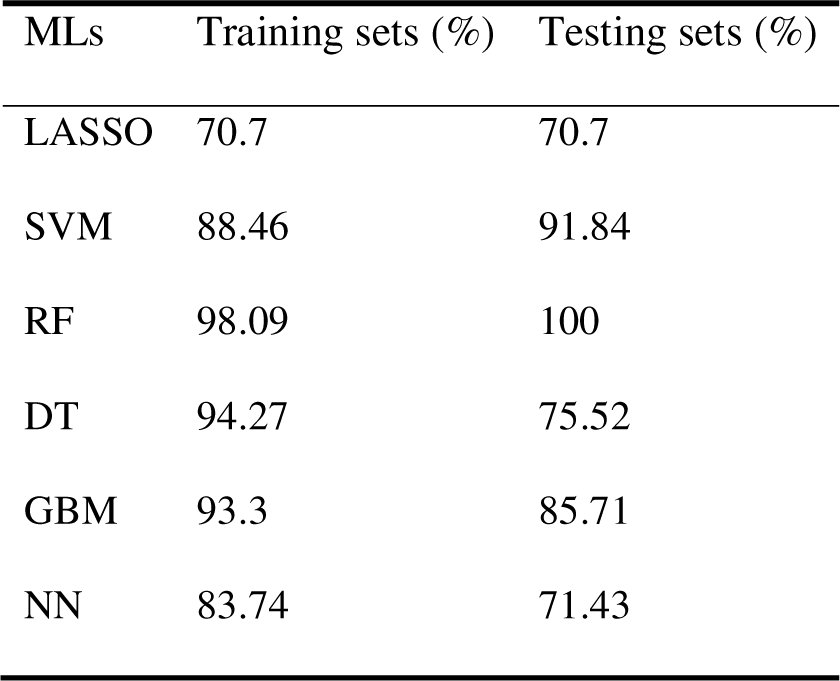
Accuracy of six MLs based on 39 DEGs in the training and test sets.

| MLs | Training sets (%) | Testing sets (%) |
| --- | --- | --- |
| LASSO | 70.7 | 70.7 |
| SVM | 88.46 | 91.84 |
| RF | 98.09 | 100 |
| DT | 94.27 | 75.52 |
| GBM | 93.3 | 85.71 |
| NN | 83.74 | 71.43 |

**Table 4.** Overall weights of six classification models were constructed to optimize the candidate diagnostic genes.

| ID | SVM | RF | NN | GBM | DT | LASSO | Overall weights |
| --- | --- | --- | --- | --- | --- | --- | --- |
| ASGR2 | 1 | 1 | 0.97 | 0.21 | 1 | 0.61 | 4.79 |
| SOCS3 | 0.98 | 0.34 | 0.52 | 0.24 | 0.59 | 0.61 | 3.28 |
| AQP9 | 0.61 | 0.1 | 0.68 | 1 | 0 | 0.52 | 2.91 |
| PPARG | 0.76 | 0.15 | 1 | 0.25 | 0 | 0.5 | 2.66 |
| RNASE1 | 0.74 | 0.22 | 0.24 | 0.1 | 0.41 | 0.74 | 2.45 |
| DYSF | 0.2 | 0.68 | 0.01 | 0.57 | 0 | 0.72 | 2.18 |
| S100A9 | 0.53 | 0.09 | 0.01 | 0.74 | 0 | 0.63 | 2 |
| FCGR1A | 0.17 | 0.51 | 0 | 0.57 | 0 | 0.68 | 1.93 |
| VSIG4 | 0.44 | 0.3 | 0.1 | 0.19 | 0.01 | 0.86 | 1.9 |
| STAB1 | 0.47 | 0.58 | 0.05 | 0.14 | 0 | 0.61 | 1.85 |
| MYBL1 | 0.68 | 0.15 | 0.2 | 0 | 0.26 | 0.52 | 1.81 |
| KLRD1 | 0.26 | 0.65 | 0.01 | 0.05 | 0 | 0.73 | 1.7 |
| ECRP | 0.44 | 0.24 | 0.11 | 0 | 0.34 | 0.54 | 1.67 |
| TCN2 | 0.46 | 0.27 | 0.07 | 0 | 0 | 0.78 | 1.58 |
| FAM20A | 0.31 | 0.08 | 0.15 | 0 | 0 | 1 | 1.54 |
| MERTK | 0.19 | 0.21 | 0.01 | 0.1 | 0.14 | 0.71 | 1.36 |
| HP | 0.09 | 0.78 | 0 | 0 | 0 | 0.45 | 1.32 |
| RNASE2 | 0.16 | 0.42 | 0.01 | 0 | 0 | 0.7 | 1.29 |
| DTHD1 | 0.13 | 0.45 | 0.05 | 0 | 0 | 0.66 | 1.29 |
| CLC | 0.11 | 0.72 | 0.02 | 0 | 0 | 0.36 | 1.21 |
| SNORD20 | 0.14 | 0.24 | 0.01 | 0.13 | 0.1 | 0.5 | 1.12 |
| CD163 | 0.15 | 0.29 | 0 | 0.11 | 0 | 0.57 | 1.12 |
| NRG1 | 0.2 | 0.25 | 0.02 | 0 | 0 | 0.63 | 1.1 |
| SNORD45B | 0.12 | 0.64 | 0.01 | 0 | 0 | 0.33 | 1.1 |
| CYP1B1 | 0.14 | 0.25 | 0 | 0 | 0 | 0.66 | 1.05 |
| KLRC2 | 0.07 | 0.51 | 0 | 0 | 0 | 0.46 | 1.04 |
| TMEM176A | 0.08 | 0.67 | 0 | 0 | 0 | 0.24 | 0.99 |
| SLED1 | 0.09 | 0.24 | 0.02 | 0.05 | 0 | 0.49 | 0.89 |
| FCGR1A.2 | 0.23 | 0 | 0 | 0.62 | 0 | 0 | 0.85 |
| SERPINB2 | 0.08 | 0.21 | 0 | 0 | 0 | 0.54 | 0.83 |
| FCGR1A.1 | 0.18 | 0 | 0 | 0.62 | 0 | 0 | 0.8 |
| KLRC4 | 0.13 | 0.21 | 0 | 0 | 0 | 0.43 | 0.77 |
| KLRA1P | 0.1 | 0.07 | 0 | 0.08 | 0 | 0.51 | 0.76 |
| MIR21 | 0.08 | 0.09 | 0.01 | 0 | 0 | 0.5 | 0.68 |
| CES1 | 0.12 | 0.05 | 0.03 | 0 | 0 | 0.47 | 0.67 |
| KLRC4-KLRK1 | 0.07 | 0 | 0 | 0.08 | 0 | 0.43 | 0.58 |
| KLRC3 | 0.07 | 0.1 | 0 | 0 | 0 | 0.39 | 0.56 |
| NRG1.1 | 0.13 | 0 | 0 | 0 | 0 | 0 | 0.13 |
| FMN1.1 | 0.07 | 0 | 0.01 | 0 | 0 | 0 | 0.08 |

With the basis of overall normalized weights >1, 26 candidate genes were filtered for subsequent diagnosis in AMI and control groups in the training and testing sets. Among the 26 genes, 10 were excluded because of no differentiation in the testing set. Sixteen genes were significant in the two sets (**Fig. 5**).

**Fig. 5.**
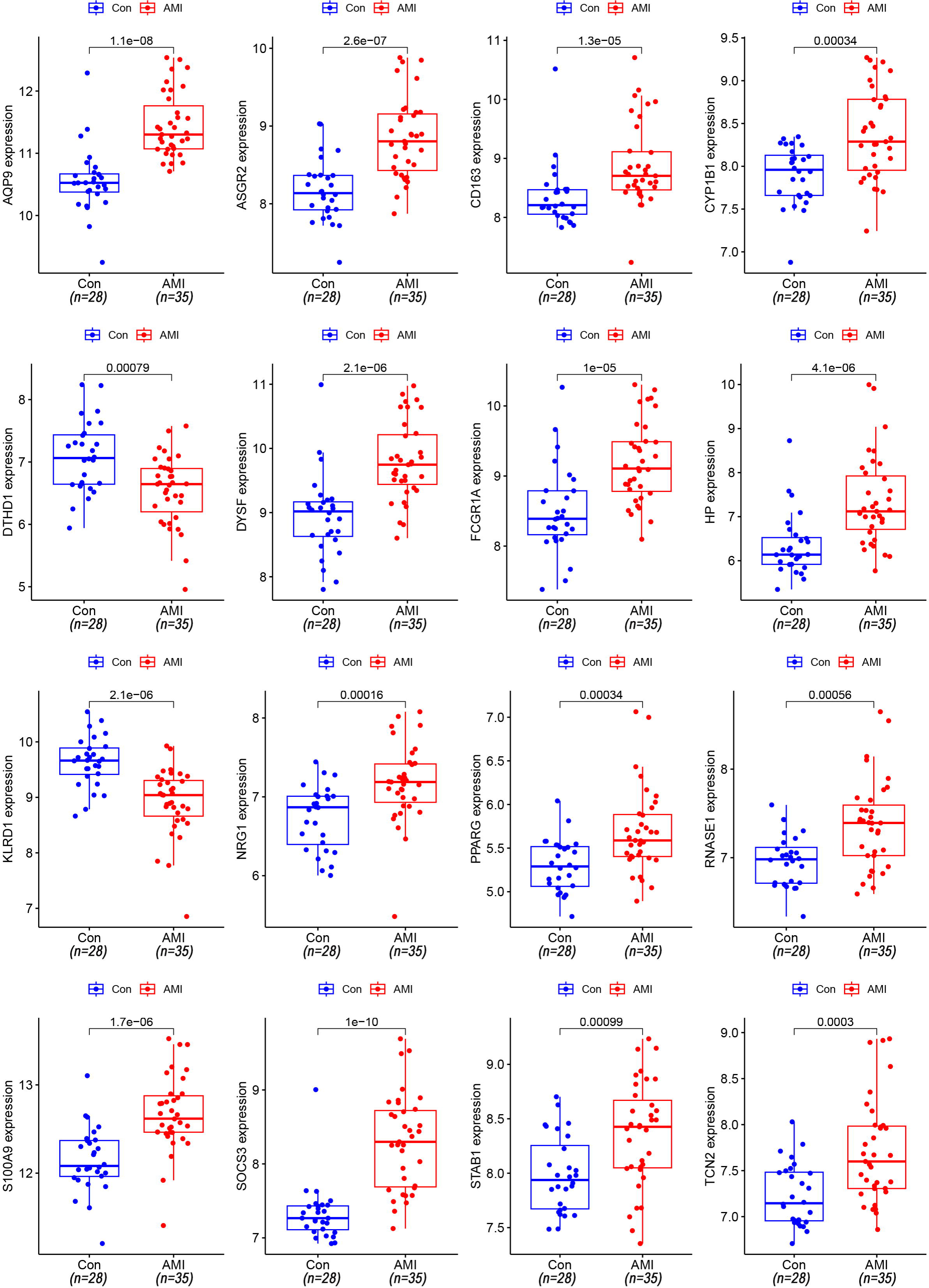
The 16 DEGs also differed in the testing set.

### 3.5 Diagnosis value of candidate genes

Sixteen candidate genes were included in the following ROC analysis. The AUC values of SOCS3, AQP9, and ASGR2 were greater than 0.85 in both the training and testing sets. In particular, 2 genes, SOCS3 and AQP9, were greater than 0.9 (**Fig. 6**). The AUC value of the two genes indicated a potential diagnostic value in AMI.

**Fig. 6.**
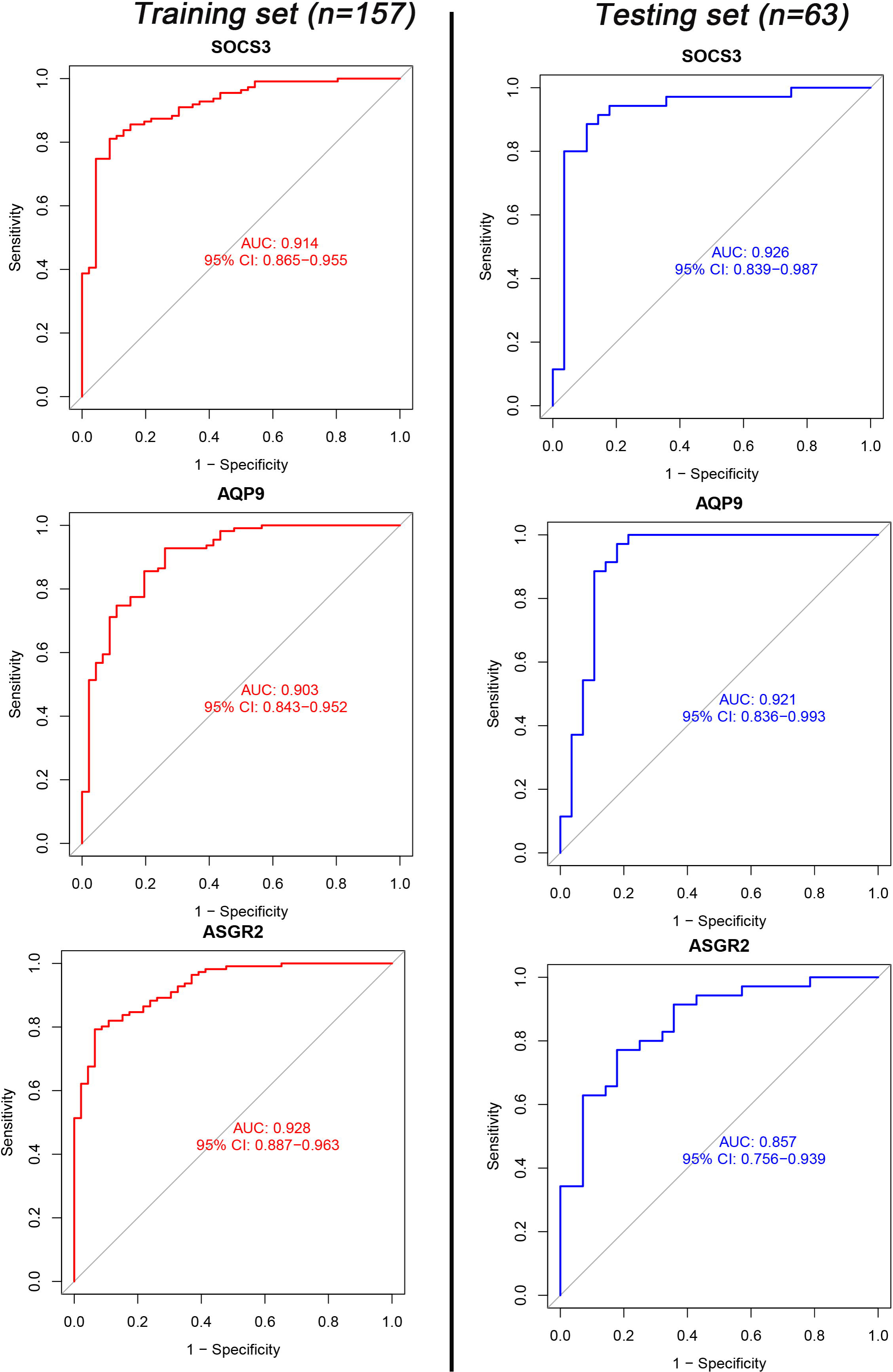
ROC curves for AQP9, SOCS3, and ASGR2 in the training and testing sets.

### 3.6 Correlation analysis

Immune correlation was performed with the 220 samples (**Fig. 7**). The infiltration landscape (**Fig. 7A**) showed 22 immune distributions in the control and AMI groups. Nine types of immune cells (CD8 T cells, naive CD4 T cells, regulatory T cells (Tregs), resting NK cells, monocytes, M0 macrophages, M2 macrophages, eosinophils, and neutrophils) infiltrated significantly between the control and AMI groups (**Fig. S1**). Moreover, the correlations between 22 immunized cells and the two diagnostic genes, AQP9 and SOCS3, based on *Spearman* analysis (**Fig. 7B-C**) showed significant correlations with 9 immune cells (monocytes, neutrophils, CD8 T cells, resting NK cells, naive CD4 T cells, eosinophils, M2 macrophages, activated dendritic cells, and memory B cells). More importantly, two immune cell types (monocytes and neutrophils) possessed a higher correlation coefficient (**Fig. 7B-C**) than the other 7 immune cell types (**Fig. S2-S3**). In particular, the correlation coefficients of monocytes (**Fig. 7B-C**) were highest for the two genes (0.56 for SOCS3 and 0.76 for AQP9).

**Fig. 7.**
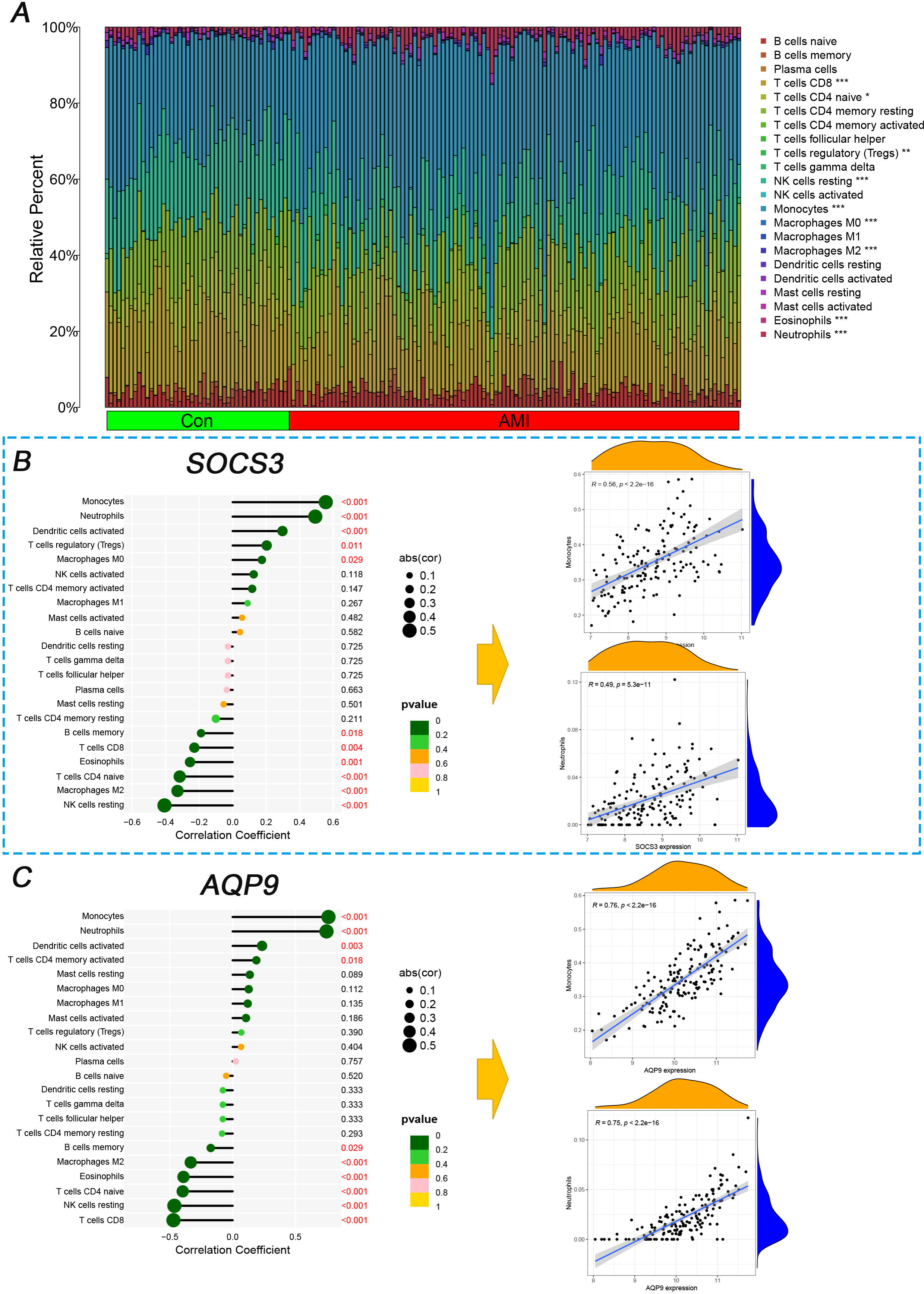
Immune correlation analysis of AQP9 and SOCS3 between the control and AMI groups. **A** The stacked column graph between the control and AMI groups. **B** The violin plot showed 7 immune cell types infiltrated differently between the control and AMI groups. **C** The lollipop map of the different immune cell types in AQP9 and SOCS3. * mean <0.05, ** mean <0.01, ***mean<0.001.

### 3.7 Clinical validation

Finally, 40 individuals (20 SCAD and 20 AMI patients) were recruited. The general information of these individuals was shown in **Table 5**. Among 39 clinical characteristics were summarized, and 13 had significance between the SCAD and AMI patients, including WBC, NeP, MonP, Lym, GAT, D-dimer, CRP, SOCS3, AQP9, LDH, cTnT, CK-MB, and Albumin.

**Table 5.** The general characteristics of the 40 patients.

| Characteristic | SCAD (*n*= 20) | AMI (*n*= 20) | *P*-value |
| --- | --- | --- | --- |
| Hypertension, % | 16.00(80) | 16.00(80) | >0.05 |
| Diabetes mellitus, % | 6.00(30) | 7.00(35) | >0.05 |
| Stroke, % | 4.00(20) | 4.00(20) | >0.05 |
| Hyperlipemia, % | 4.00(20) | 6.00(30) | >0.05 |
| Age, year | 66 (63, 72) | 70 (58, 77) | >0.05 |
| Sex (Male), % | 10.00(50.00) | 12.00(60.00) | >0.05 |
| RBC, million cells/μL | 4.37 (3.80, 4.49) | 4.03 (3.55, 4.35) | >0.05 |
| WBC, 1000 cells/μL | 6.02 (4.85, 6.84) | 8.33 (6.84, 11.24) | <0.001 |
| NeP, % | 69 (58, 74) | 80 (75, 86) | <0.001 |
| MonP, % | 8.00 (6.15, 9.23) | 5.55 (3.98, 7.68) | <0.05 |
| Mon, 1000 cells/μL | 0.44 (0.35, 0.48) | 0.42 (0.19, 0.73) | >0.05 |
| Lym, 1000 cells/μL | 1.37 (1.06, 1.77) | 0.96 (0.62, 1.42) | <0.05 |
| RDW, % | 13.00 (12.55, 13.30) | 13.55 (12.88, 14.98) | >0.05 |
| PDW, % | 11.85 (10.48, 13.90) | 13.25 (11.68, 16.15) | >0.05 |
| Pla, 1000 cells/μL | 214 (163, 245) | 219 (173, 244) | >0.05 |
| MCHC, g/L | 334 (329, 342) | 329 (319, 338) | >0.05 |
| Hg, g/L | 131 (113, 140) | 119 (109, 134) | >0.05 |
| GAT, U/L | 16 (14, 21) | 28 (17, 51) | <0.05 |
| D-dimer, mg/L | 0.46 (0.27, 0.69) | 1.01 (0.62, 2.70) | <0.01 |
| CRP, mg/L | 1 (1, 2) | 12 (7, 26) | <0.001 |
| SOCS3 | 1.57 (1.22, 1.76) | 1.97 (1.86, 2.20) | <0.001 |
| AQP9 | 0.90 (0.85, 1.03) | 1.44 (1.16, 1.66) | <0.001 |
| LDH, U/L | 152 (141, 194) | 260 (228, 303) | <0.001 |
| cTnT, μg/mL | 12 (9, 18) | 140 (92, 264) | <0.001 |
| CK-MB, U/L | 2 (1, 4) | 19 (9, 33) | <0.001 |
| LDL, mmol/L | 1.83 (1.57, 2.68) | 2.38 (1.74, 3.62) | >0.05 |
| HDL, mmol/L | 1.04 (0.96, 1.16) | 1.11 (0.96, 1.34) | >0.05 |
| TC, mmol/L | 3.59 (2.87, 4.52) | 4.15 (3.27, 5.80) | >0.05 |
| TG, mmol/L | 1.13 (0.71, 1.58) | 0.98 (0.89, 1.22) | >0.05 |
| Glucose, mg/L | 5.36 (4.73, 5.81) | 6.05 (5.12, 9.15) | >0.05 |
| Cys, μmol/L | 11.7 (10.1, 16.4) | 14.2 (8.7, 22.1) | >0.05 |
| Albumin, g/L | 41.8 (38.9, 43.3) | 38.6 (34.9, 40.3) | <0.01 |
| Total protein, g/L | 65 (63, 67) | 65 (60, 67) | >0.05 |
| GGT, U/L | 13 (11, 22) | 17 (12, 33) | >0.05 |
| IBIL, μmol/L | 3.65 (2.50, 6.03) | 4.95 (3.50, 7.30) | >0.05 |
| DBIL, μmol/L | 4.00 (2.58, 4.78) | 4.30 (2.85, 7.75) | >0.05 |
| IBIL, μmol/L | 7.8 (5.2, 10.7) | 10.0 (6.3, 17.4) | >0.05 |
| Globulin, g/L | 24.7 (21.2, 25.8) | 25.6 (23.5, 26.9) | >0.05 |
| ALP, U/L | 72 (61, 87) | 87 (72, 108) | >0.05 |

RBC; red blood cell count, WBC; white blood cell count, NeP; neutrophils percentage, MonP; monocyte percentage, Mon; monocyte count, Lym; lymphocyte count, RDW; red blood cell distribution width, PDW; platelet distribution width, Pla; platelet count, MCHC; mean corpuscular haemoglobin concentration, Hg; haemoglobin, GAT; glutamic transaminase, CRP; c-reactive protein, LDH; lactate dehydrogenase, cTnT; cardiac troponin t, CK-MB; creatine kinase isoenzymes, LDL; low-density lipoprotein, HDL; high-density lipoprotein, TC; total cholesterol, TG; total triglycerides, Cys; homocysteine, GGT; gammaglutaminase, IBIL; indirect bilirubin, DBIL; direct bilirubin, IBIL; total bile acid, ALP; alkaline phosphatase.

The relative RNA levels (**Fig. 8A**) of AQP9 and SOCS3 were both significant. The SOCS3 content of coronary arteries differed by the number of lesions (**Fig. 8B**): three lesions showed significantly higher SOCS3 than two and one (**Fig. 8B**). Patients with III-IV Killip classification had higher SOCS3 compared to those with I-II (**Fig. 8C**). Although more stenotic coronary arteries were associated with higher levels of AQP9, the difference was less significant than for SOCS3 (**Fig. 8B**). In addition, different Killip classifications associated with AQP9 possessed no significant differences (**Fig. 8C**). Furthermore, the 9 significant clinical features were analysised with *Pearson* correlation test (**Fig. S4**). And SOCS3 had a positive correlation with AQP9. Both genes had a negative correlation with Albumin.

**Fig. 8.**
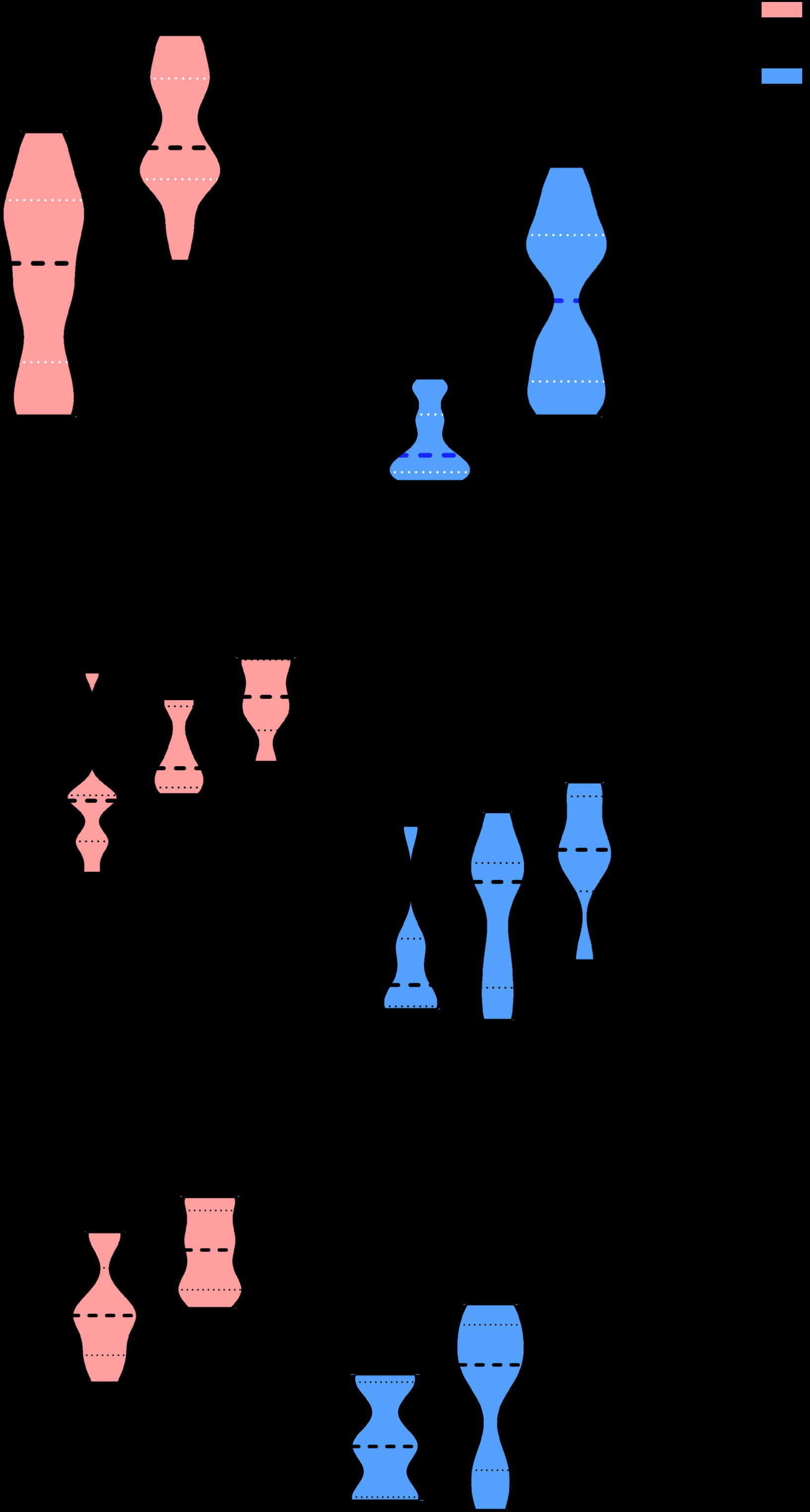
Relative RNA levels of AQP9 and SOCS3 in AMI patients and SCAD controls. **A** The relative content of SOCS3 and AQP9 in AMI patients and SCAD controls. **B** The comparison of AQP9 and SOCS3 in the number of coronary arteries with different stenoses in AMI. **C** The comparison of AQP9 and SOCS3 in various Killip classifications in AMI. * mean <0.05, ** mean <0.01, *** mean<0.001, ns mean no significance.

## 4 Discussion

To our knowledge, our work is the first to filter AMI diagnosis genes based on the overall normalized weights of IML. Four microarrays with 220 samples were adopted for data analysis, and further clinical studies were performed to validate the results. Two genes, AQP9 and SOCS3, showed an AUC >0.9 in both the training set and testing set (**Fig. 6**). Both genes showed a typical and highest correlation coefficient (**Fig. S2-3**) in monocytes. The clinical study verified the significance between AMI patients and healthy controls, indicating a potential diagnostic value of AQP9 and SOCS3. Compared with previous studies, we reached similar conclusions that AQP9 presented diagnostic value for AMI [40, 41], and we further explored the immune correlation of AQP9. Additionally, Prof. Zhu [42] identified SOCS3 as an immune-related gene in AMI, and we expanded it to have diagnostic value. More importantly, this study is the first to reveal the RNA correlation of AQP9 and SOCS3, especially SOCS3, between the number of stenotic coronary arteries and the Killip classification.

AQP9, a cell membrane protein, transports water down the concentration gradient. ERK1/2 can be reversed in AMI rats by silencing AQP9, attenuating cardiomyocytes’ inflammatory response and apoptosis and upregulating cardiac function [43]. The above research indicated the crucial role of AQP9 in the pathogenesis of AMI. In human polymorphonuclear leukocytes, AQP9-related inflammation may result from the NK-κB [44] and F-actin polymerization [45]. In our work, the ROC curve of AQP9 was > 0.9. Therefore, AQP9 might be a potential genetic marker for diagnosing AMI with SCAD.

SOCS3 is increased in AMI mice [29] and regulates the T-cell repertoire with STAT3/SOCS3 signalling [46]. More importantly, cardiac-specific silencing of SOCS3 triggers sustained STAT3 and decreases myocardial apoptosis [47]. Therefore, SOCS3 is the dominant negative modulator [48] of Th17 via STAT3 [49]. Apoptosis regulates the pathophysiological evaluation of AMI [50]. In vitro, SOCS3 can trigger the apoptosis of mammary cells [51], and knocking out SOCS3 regulates the expression of apoptosis in 3T3-L1 preadipocytes [52]. The above research emphasized the immune regulation of SOCS3 and the regulation of apoptosis with STAT3. In our work, the ROC curve of SOCS3 was > 0.9. Therefore, SOCS3 might be an effective genetic marker for diagnosing AMI.

Additionally, the CIBERSORT algorithm showed that the proportion of neutrophils and monocytes in the AMI group was higher than in the control group. The progression of AMI is correlated with immune disorder. For example, the white blood cell count correlates highly with in-hospital mortality after AMI [53]. Neutrophils are increased in peripheral blood, and researchers have emphasized that neutrophils-lymphocytes [54, 55] and monocytes/macrophages [56] can be easily acquired factors for the prognosis of AMI. Macrophages were dominant in infarcted myocardium, especially over the first week of AMI [57]. However, NK cells have diminished cytotoxic function [58], and the targeted regulation of NK cells may indicate a dominant role in the cure of AMI. At the beginning of AMI, inflammation deteriorates with increased neutrophils and monocytes [59], and inflammation decreases over time with the reduced function of NK cells. Innate immunity is a vital regulatory factor in the inflammatory, proliferative, and maturation phases [3, 60, 61]. AMI leads to a deteriorated inflammatory process. Currently, novel therapeutic interventions targeting the immune system may regulate slant inflammation, which is conducive to resolving pathological conditions. In a previous clinical trial of 182 NSTEMI patients (a subtype of AMI), the patient’s intake of IL-1 blockers decreased acute inflammation [62]. Another immune study showed that short-term blockade of S100A9 downregulates inflammation [63] in permanent coronary ischaemia mice. However, the above immune interventions are still experimental and not in the clinic. In summary, regulating immune cells along with the progression of AMI and immune intervention in AMI might be a potential target.

AQP9 expression was highest in human polymorphonuclear leukocytes [45] compared with the spleen and liver, suggesting a possible correlation between AQP9 and immunity or inflammation. AQP9 regulates water flow on leukocytes [64], which regulates cellular morphology and motility, a change that facilitates the migration of leukocytes to inflammatory sites. Similar to our result, Hawang [65] indicated the correlation between AQP9 and neutrophile granulocytes. Research [29, 60, 61] emphasizes the correlation between SOCS3 and neutrophils in inflammation. In our research, both genes had a higher correlation with two immune cells, neutrophils and monocytes. The immune cell correlation indicated that the targeted gene therapy of immune cells may benefit the course of AMI—potential feasibility of using AQP9 and SOCS3 as therapeutic targets or predictors of treatment response.

ML algorithms are widely performed for various cardiovascular diseases, such as optimizing variables, classification, and congression. For variable filtration, numerous studies take only single or double ML algorithms (e.g., weighted gene coexpression network analysis [66], LASSO, and SVM). However, only the single or double ML algorithms might unconsciously delete the potential genes. For example, AQP9 will be ignored if we only take DT because the weights of AQP9 were zero in DT (**Table 4**). Taking only a single ML might miss some potential genes. For example, although LASSO can detect candidate genes with big data when highly correlated features exist, the LASSO regression method tends to select one of them and ignore all the other features, leading to the instability of the results [67]. In pigmented skin lesions [68], SVM and NN displayed their talent classification value. In preoperative postsurgical mortality [69], GBM is optimized rather than DT, RF, and SVM. Various ML algorithms may show different weights even in the same variable (**Table 4**). Necessarily, the overall normalized weights of IML were taken to filter genes. Surprisingly, IML explores two potential, unreported diagnostic genes in AMI. In our study, IML has good value in both variable screening and model prediction.

Inevitably, three limitations exist in this work, although the best efforts were taken to eliminate them. Primarily, small sample size verification might possess some bias. So, multicentre collaborations or leveraging larger external datasets is crutial for further verification. Although testing sets and clinical validation were developed to assess the stability of the diagnostic value, the bias of single-centre validation might exist. More confirmation, clinical trials and animal experiments are indispensable for solid verification. Next, the ML algorithms contained limitations (e.g., the black box phenomenon [70]), especially NN, which has numerous layers [71]. The set of operations an ML performs in making a prediction is unknown, even if a human knows precisely what the model is doing at each step of the decision-making process. The operations performed cannot be described in terms of human-understandable semantics. And the Interpretability techniques for ML models always catch the eye of developers, which enhances the transparency and reliability of the ML. Finally, limited clinical features were obtained (e.g., age [72], ethnicity, and race [73]). Clinical features could potentially enhance the predictive accuracy of the diagnostic model and provide a more comprehensive understanding of AMI. For example, various combinations (*e.g.,* sex, smoking or not, and laboratory indicators) of clinical variables [74] are calibrated to analyze the relationship between the target variable and the outcome.

## 5 Conclusion

Based on the overall normalized weights of IML, the research successfully merges four microarrays and uncovers hidden diagnostic genes AQP9 and SOCS3 for leukocytes of AMI patients. AQP9 and SOCS3 are closely associated with monocytes and neutrophils, which might contribute to advancing AMI diagnosis and shedding light on novel genetic markers, including AMI pathogenesis, targeted therapies, and potential precision medicine. Although clinical validation copies the result again. Multiple clinical characteristics, multicenter, and large-sample relevant trials are still needed to confirm its clinical value.

## Authors’ contributions

LZ and YL wrote the original draft. LZ, KYW, YL, JSZ, and HLZ performed the research. LZ, YL, XZ, and KYW analyzed the data. SQ, MH, JSZ, and HLZ designed the experiment and revised the manuscript.

## Founding

The research was funded by Suzhou Science & Technology Development Plan (SYSD2019222). Zhangjiagang science and technology plan project (ZKS2135), Youth science and technology project of Zhangjiagang Municipal Health Commission (ZJGQNKJ202211).

## Consent for publication

This study has not been published before, and this publication has been approved by all authors.

## Ethics approval and consent to participate

The clinical trial part was approved by the Ethics Review Committee of Jinghai District Hospital.

## Availability of data and material

The datasets presented in this study can be found online. The names of the repositories and GEO numbers can be found below: https://www.ncbi.nlm.nih.gov/geo/query/acc.cgi?acc=GSE59867;https://www.ncbi.nlm.nih.gov/geo/query/acc.cgi?acc=GSE60993;https://www.ncbi.nlm.nih.gov/geo/query/acc.cgi?acc=GSE62646;https://www.ncbi.nlm.nih.gov/geo/query/acc.cgi?acc=GSE48060.

## Abbreviation

AUC: Area under the Curve
AMI: Acute Myocardial Infarction
IML: Integration Machine Learning
DEGs: Differently Expressed Genes
KEGG-GSEA: Kyoto Encyclopedia of Genes and Genomes-Gene Set Enrichment Analysis
GO: Gene Ontology
DO: Disease Ontology
MF: Molecular Function
BP: Biological Process
CC: Cellular Components
SVM: Support Vector Machine
ML: Machine Learning
LASSO: Least Absolute Shrinkage and Selection Operator
RF: Random Forest
GBM: Gradient Boosting Machine
DT: Decision Trees
NN: Neural Network.

## Acknowledgments

We thank Suzhou Science & Technology Development Plan.

