## Supplementary material for "Integration of Machine Learning to Identify Diagnostic Genes in Leukocytes for Acute Myocardial Infarction Patients": Table S1, Table S2, Table S3, Table S4, Table S5, Fig S1, Fig S2, Fig S3, Fig S4.

**Content**

Table S1. The 39 DEGs in healthy and AMI.

Table S2. GSEA enrichment of 45 terms.

Table S3. GO enrichment of 160 terms.

Table S4. DO enrichment of 57 terms.

Table S5. Primary weight of DEGs in the six classifications MLs.

Fig.S1 The difference between the 22 immune cells.

Fig S2. The other 7 immune cells correlation analysis of SOCS3.

Fig S3. The other 7 immune cells correlation analysis of AQP9.

Fig S4. The Correlation analysis of 9 clinical variables.

Table S1. The 39 DEGs in healthy and AMI.

| Genes | logFC | AveExpr | t | P.Value | adj.P.Val | B |
| --- | --- | --- | --- | --- | --- | --- |
| ASGR2 | 0.8 | 8.8 | 11.2 | 5.98E-22 | 3.24E-18 | 39.2 |
| AQP9 | 1 | 10.2 | 10.7 | 1.64E-20 | 5.16E-17 | 36 |
| SOCS3 | 1.2 | 8.6 | 10.6 | 4.16E-20 | 1.05E-16 | 35.1 |
| S100A9 | 0.7 | 11.2 | 10.5 | 4.36E-20 | 1.05E-16 | 35 |
| STAB1 | 0.9 | 9.6 | 10.2 | 3.20E-19 | 4.62E-16 | 33.1 |
| ECRP | 1.1 | 8.8 | 9.9 | 1.95E-18 | 2.01E-15 | 31.3 |
| MYBL1 | -0.8 | 9.8 | -9.3 | 1.01E-16 | 4.65E-14 | 27.5 |
| DYSF | 0.8 | 8.7 | 9 | 5.92E-16 | 1.78E-13 | 25.7 |
| CYP1B1 | 0.8 | 9.8 | 8.6 | 6.43E-15 | 1.14E-12 | 23.4 |
| RNASE1 | 0.8 | 7.7 | 8.3 | 3.60E-14 | 4.31E-12 | 21.7 |
| RNASE2 | 0.8 | 9.7 | 8.3 | 4.39E-14 | 4.95E-12 | 21.5 |
| PPARG | 0.8 | 6.1 | 8.3 | 4.43E-14 | 4.98E-12 | 21.5 |
| FCGR1A.2 | 0.8 | 9.3 | 8 | 1.88E-13 | 1.52E-11 | 20.1 |
| TCN2 | 0.8 | 8.2 | 8 | 2.17E-13 | 1.69E-11 | 20 |
| MERTK | 0.8 | 8.8 | 7.9 | 3.74E-13 | 2.69E-11 | 19.5 |
| FCGR1A.1 | 0.8 | 9.3 | 7.8 | 5.66E-13 | 3.73E-11 | 19.1 |
| KLRD1 | -0.7 | 9.9 | -7.8 | 6.61E-13 | 4.24E-11 | 18.9 |
| MIR21 | 0.8 | 10.4 | 7.5 | 3.60E-12 | 1.65E-10 | 17.3 |
| FCGR1A | 0.8 | 9.7 | 7.2 | 2.40E-11 | 7.59E-10 | 15.4 |
| CD163 | 0.7 | 8.8 | 7.1 | 3.08E-11 | 9.43E-10 | 15.2 |
| DTHD1 | -0.7 | 7.8 | -7.1 | 3.56E-11 | 1.06E-09 | 15 |
| FAM20A | 1 | 6.9 | 7 | 5.70E-11 | 1.56E-09 | 14.6 |
| HP | 1.1 | 7 | 7 | 6.54E-11 | 1.77E-09 | 14.4 |
| NRG1 | 0.8 | 7.6 | 7 | 6.92E-11 | 1.84E-09 | 14.4 |
| SERPINB2 | 0.7 | 7.9 | 6.9 | 1.04E-10 | 2.56E-09 | 14 |
| VSIG4 | 0.8 | 7.6 | 6.9 | 1.07E-10 | 2.62E-09 | 14 |
| KLRA1P | -0.7 | 7.4 | -6.9 | 1.25E-10 | 2.99E-09 | 13.8 |
| CES1 | 0.9 | 9 | 6.7 | 3.64E-10 | 7.44E-09 | 12.8 |
| KLRC4 | -0.8 | 9.4 | -6.6 | 6.21E-10 | 1.17E-08 | 12.3 |
| SNORD20 | -0.8 | 6.1 | -6.4 | 1.98E-09 | 3.14E-08 | 11.1 |
| NRG1.1 | 0.8 | 7.1 | 6.2 | 4.58E-09 | 6.46E-08 | 10.3 |
| SLED1 | 0.7 | 8.2 | 6.1 | 9.12E-09 | 1.15E-07 | 9.7 |
| FMN1.1 | 0.7 | 7.2 | 6 | 1.12E-08 | 1.37E-07 | 9.5 |
| SNORD45B | -0.9 | 7.6 | -6 | 1.17E-08 | 1.42E-07 | 9.4 |
| KLRC4-KLRK1 | -0.8 | 9.5 | -6 | 1.58E-08 | 1.85E-07 | 9.1 |
| KLRC2 | -0.8 | 6.4 | -5.7 | 5.89E-08 | 5.83E-07 | 7.9 |
| KLRC3 | -0.7 | 8 | -5.4 | 2.03E-07 | 1.73E-06 | 6.7 |
| CLC | -0.8 | 7.3 | -4.3 | 3.46E-05 | 0.000150984 | 1.8 |
| TMEM176A | 0.7 | 9.3 | 3.2 | 0.001914651 | 0.004857423 | -1.9 |

* logFC, the expression value of of fold change; AveExpr, the average expression of genes; adj.P.Val, the adjusted value of P.

Table S2. GSEA enrichment of 45 terms.

| ID | setSize | enrichmentScore | NES | pvalue | p.adjust |
| --- | --- | --- | --- | --- | --- |
| Lysosome | 121 | 0.6 | 2.5 | 1.00E-10 | 1.84E-08 |
| Fc_Gamma_R_Mediated_Phagocytosis | 94 | 0.5 | 2 | 4.57E-06 | 0.00035 |
| Huntingtons_Disease | 156 | 0.4 | 1.9 | 5.71E-06 | 0.00035 |
| Ppar_Signaling_Pathway | 68 | 0.5 | 2.1 | 8.39E-06 | 0.000386 |
| Leishmania_Infection | 65 | 0.5 | 2 | 7.52E-05 | 0.001976 |
| Oxidative_Phosphorylation | 99 | 0.4 | 1.8 | 5.40E-05 | 0.001976 |
| Natural_Killer_Cell_Mediated_Cytotoxicity | 124 | -0.4 | -1.8 | 7.20E-05 | 0.001976 |
| Glutathione_Metabolism | 50 | 0.5 | 2 | 8.99E-05 | 0.002067 |
| Glycolysis_Gluconeogenesis | 61 | 0.5 | 1.9 | 1.33E-04 | 0.002439 |
| Cell_Cycle | 122 | -0.4 | -1.8 | 1.23E-04 | 0.002439 |
| T_Cell_Receptor_Signaling_Pathway | 108 | -0.4 | -1.8 | 1.86E-04 | 0.003112 |
| Complement_And_Coagulation_Cascades | 68 | 0.5 | 1.9 | 2.75E-04 | 0.003893 |
| Chemokine_Signaling_Pathway | 184 | 0.3 | 1.6 | 2.67E-04 | 0.003893 |
| Alzheimers_Disease | 143 | 0.4 | 1.7 | 3.66E-04 | 0.004816 |
| Bladder_Cancer | 41 | 0.5 | 1.8 | 5.47E-04 | 0.006712 |
| Prion_Diseases | 35 | 0.5 | 1.8 | 7.47E-04 | 0.00859 |
| Drug_Metabolism_Other_Enzymes | 40 | 0.5 | 1.9 | 8.49E-04 | 0.009186 |
| Nod_Like_Receptor_Signaling_Pathway | 62 | 0.5 | 1.8 | 9.54E-04 | 0.009234 |
| Mapk_Signaling_Pathway | 263 | 0.3 | 1.5 | 9.37E-04 | 0.009234 |
| Hematopoietic_Cell_Lineage | 83 | 0.4 | 1.7 | 1.33E-03 | 0.012252 |
| Endocytosis | 175 | 0.3 | 1.6 | 1.52E-03 | 0.013302 |
| Spliceosome | 123 | -0.4 | -1.6 | 1.67E-03 | 0.014003 |
| Other_Glycan_Degradation | 16 | 0.7 | 1.9 | 1.76E-03 | 0.01411 |
| Acute_Myeloid_Leukemia | 56 | 0.5 | 1.8 | 2.01E-03 | 0.014769 |
| Adipocytokine_Signaling_Pathway | 66 | 0.4 | 1.7 | 1.95E-03 | 0.014769 |
| Insulin_Signaling_Pathway | 136 | 0.3 | 1.5 | 2.38E-03 | 0.016874 |
| Metabolism_Of_Xenobiotics_By_Cytochrome_P450 | 61 | 0.4 | 1.7 | 2.55E-03 | 0.017374 |
| Drug_Metabolism_Cytochrome_P450 | 61 | 0.4 | 1.7 | 2.73E-03 | 0.017909 |
| Galactose_Metabolism | 25 | 0.6 | 1.9 | 3.69E-03 | 0.021897 |
| Antigen_Processing_And_Presentation | 67 | -0.4 | -1.7 | 3.50E-03 | 0.021897 |
| Ubiquitin_Mediated_Proteolysis | 129 | -0.3 | -1.5 | 3.65E-03 | 0.021897 |
| Rna_Degradation | 55 | -0.4 | -1.7 | 3.94E-03 | 0.022656 |
| Glycosphingolipid_Biosynthesis_Globo_Series | 14 | 0.7 | 1.8 | 4.89E-03 | 0.027277 |
| Maturity_Onset_Diabetes_Of_The_Young | 24 | 0.6 | 1.8 | 5.34E-03 | 0.028056 |
| Taste_Transduction | 51 | -0.5 | -1.7 | 5.22E-03 | 0.028056 |
| Glycerophospholipid_Metabolism | 75 | 0.4 | 1.6 | 5.59E-03 | 0.028597 |
| Toll_Like_Receptor_Signaling_Pathway | 99 | 0.3 | 1.5 | 5.83E-03 | 0.028994 |
| Arachidonic_Acid_Metabolism | 56 | 0.4 | 1.6 | 0.006278 | 0.030397 |
| Graft_Versus_Host_Disease | 29 | -0.5 | -1.7 | 0.006891 | 0.032406 |
| Cytokine_Cytokine_Receptor_Interaction | 256 | 0.3 | 1.4 | 0.007045 | 0.032406 |
| Fructose_And_Mannose_Metabolism | 32 | 0.5 | 1.7 | 0.007285 | 0.032694 |
| Dna_Replication | 36 | -0.5 | -1.7 | 0.009766 | 0.042784 |
| Ecm_Receptor_Interaction | 82 | 0.4 | 1.5 | 0.010157 | 0.043463 |
| Protein_Export | 23 | -0.5 | -1.6 | 0.011469 | 0.047056 |
| Neuroactive_Ligand_Receptor_Interaction | 271 | 0.3 | 1.4 | 0.011508 | 0.047056 |

Table S3. GO enrichment of 160 terms.

| ONTOLOGY | Description | GeneRatio | pvalue | p.adjust |
| --- | --- | --- | --- | --- |
| BP | stimulatory C-type lectin receptor signaling pathway | 5/32 | 2.59E-10 | 1.06E-07 |
| BP | response to lectin | 5/32 | 2.59E-10 | 1.06E-07 |
| BP | cellular response to lectin | 5/32 | 2.59E-10 | 1.06E-07 |
| BP | positive regulation of natural killer cell mediated cytotoxicity | 5/32 | 7.88E-10 | 2.42E-07 |
| BP | innate immune response activating cell surface receptor signaling pathway | 5/32 | 1.65E-09 | 3.42E-07 |
| BP | positive regulation of natural killer cell mediated immunity | 5/32 | 1.95E-09 | 3.42E-07 |
| BP | innate immune response-activating signal transduction | 5/32 | 1.95E-09 | 3.42E-07 |
| BP | regulation of innate immune response | 8/32 | 4.86E-09 | 7.45E-07 |
| BP | regulation of immune effector process | 9/32 | 8.89E-09 | 1.21E-06 |
| BP | regulation of natural killer cell mediated cytotoxicity | 5/32 | 1.46E-08 | 1.79E-06 |
| BP | regulation of lymphocyte mediated immunity | 7/32 | 1.67E-08 | 1.87E-06 |
| BP | regulation of natural killer cell mediated immunity | 5/32 | 2.23E-08 | 2.28E-06 |
| BP | positive regulation of defense response | 8/32 | 3.31E-08 | 3.13E-06 |
| BP | positive regulation of lymphocyte mediated immunity | 6/32 | 4.35E-08 | 3.81E-06 |
| BP | positive regulation of leukocyte mediated cytotoxicity | 5/32 | 4.70E-08 | 3.85E-06 |
| BP | activation of innate immune response | 5/32 | 7.12E-08 | 5.46E-06 |
| BP | positive regulation of cell killing | 5/32 | 8.32E-08 | 6.01E-06 |
| BP | leukocyte mediated cytotoxicity | 6/32 | 8.83E-08 | 6.02E-06 |
| BP | positive regulation of leukocyte mediated immunity | 6/32 | 1.15E-07 | 7.08E-06 |
| BP | regulation of response to biotic stimulus | 8/32 | 1.15E-07 | 7.08E-06 |
| BP | regulation of leukocyte mediated immunity | 7/32 | 1.29E-07 | 7.52E-06 |
| BP | natural killer cell mediated cytotoxicity | 5/32 | 1.49E-07 | 8.29E-06 |
| BP | natural killer cell mediated immunity | 5/32 | 1.82E-07 | 9.69E-06 |
| BP | positive regulation of immune effector process | 7/32 | 2.21E-07 | 1.13E-05 |
| BP | regulation of leukocyte mediated cytotoxicity | 5/32 | 3.36E-07 | 1.65E-05 |
| BP | natural killer cell activation | 5/32 | 6.41E-07 | 3.02E-05 |
| BP | cell killing | 6/32 | 6.74E-07 | 3.06E-05 |
| BP | regulation of cell killing | 5/32 | 7.43E-07 | 3.26E-05 |
| BP | positive regulation of response to external stimulus | 8/32 | 7.78E-07 | 3.29E-05 |
| BP | lymphocyte mediated immunity | 7/32 | 2.18E-06 | 8.90E-05 |
| BP | positive regulation of innate immune response | 5/32 | 4.22E-06 | 0.000167 |
| BP | negative regulation of response to external stimulus | 7/32 | 6.76E-06 | 0.000259 |
| BP | negative regulation of immune system process | 7/32 | 8.19E-06 | 0.000303 |
| BP | negative regulation of innate immune response | 4/32 | 8.39E-06 | 0.000303 |
| BP | leukocyte mediated immunity | 7/32 | 1.00E-05 | 0.000351 |
| BP | positive regulation of response to biotic stimulus | 5/32 | 1.49E-05 | 0.000509 |
| BP | immune response-regulating cell surface receptor signaling pathway | 6/32 | 2.01E-05 | 0.000667 |
| BP | defense response to bacterium | 6/32 | 3.80E-05 | 0.001228 |
| BP | negative regulation of response to biotic stimulus | 4/32 | 4.24E-05 | 0.001334 |
| BP | activation of immune response | 6/32 | 4.71E-05 | 0.001446 |
| BP | regulation of natural killer cell activation | 3/32 | 4.84E-05 | 0.001448 |
| BP | response to toxic substance | 5/32 | 5.85E-05 | 0.001708 |
| BP | negative regulation of defense response | 5/32 | 8.71E-05 | 0.002486 |
| BP | negative regulation of cholesterol storage | 2/32 | 0.000151 | 0.004218 |
| BP | immune response-activating cell surface receptor signaling pathway | 5/32 | 0.000166 | 0.00442 |
| BP | immune response-activating signal transduction | 5/32 | 0.000166 | 0.00442 |
| BP | regulation of adaptive immune response based on somatic recombination of immune receptors built from immunoglobulin superfamily domains | 4/32 | 0.000242 | 0.006322 |
| BP | negative regulation of immune response | 4/32 | 0.000285 | 0.007293 |
| BP | regulation of adaptive immune response | 4/32 | 0.000334 | 0.008357 |
| BP | negative regulation of cell migration | 5/32 | 0.000355 | 0.008604 |
| BP | inflammatory response to wounding | 2/32 | 0.000372 | 0.008604 |
| BP | vascular associated smooth muscle cell apoptotic process | 2/32 | 0.000372 | 0.008604 |
| BP | regulation of vascular associated smooth muscle cell apoptotic process | 2/32 | 0.000372 | 0.008604 |
| BP | negative regulation of cell motility | 5/32 | 0.000427 | 0.009699 |
| BP | regulation of cholesterol storage | 2/32 | 0.000466 | 0.010403 |
| BP | cholesterol storage | 2/32 | 0.000571 | 0.01252 |
| BP | regulation of inflammatory response | 5/32 | 0.000608 | 0.012917 |
| BP | negative regulation of lipid storage | 2/32 | 0.000628 | 0.012917 |
| BP | cellular response to low-density lipoprotein particle stimulus | 2/32 | 0.000628 | 0.012917 |
| BP | cellular oxidant detoxification | 3/32 | 0.000637 | 0.012917 |
| BP | negative regulation of locomotion | 5/32 | 0.000642 | 0.012917 |
| BP | regulation of phagocytosis | 3/32 | 0.000695 | 0.013748 |
| BP | wound healing | 5/32 | 0.000817 | 0.015914 |
| BP | acute inflammatory response | 3/32 | 0.000933 | 0.017519 |
| BP | positive regulation of cholesterol efflux | 2/32 | 0.000949 | 0.017519 |
| BP | regulation of receptor signaling pathway via STAT | 3/32 | 0.000957 | 0.017519 |
| BP | cellular detoxification | 3/32 | 0.000957 | 0.017519 |
| BP | cellular response to toxic substance | 3/32 | 0.001162 | 0.020963 |
| BP | activation of protein kinase B activity | 2/32 | 0.001252 | 0.022264 |
| BP | smooth muscle cell apoptotic process | 2/32 | 0.001334 | 0.022968 |
| BP | regulation of smooth muscle cell apoptotic process | 2/32 | 0.001334 | 0.022968 |
| BP | response to hydrogen peroxide | 3/32 | 0.001362 | 0.022968 |
| BP | positive regulation of kinase activity | 5/32 | 0.001366 | 0.022968 |
| BP | negative regulation of receptor signaling pathway via STAT | 2/32 | 0.001419 | 0.023523 |
| BP | response to lipoprotein particle | 2/32 | 0.001506 | 0.024634 |
| BP | regulation of cellular ketone metabolic process | 3/32 | 0.001551 | 0.025039 |
| BP | negative regulation of oxidoreductase activity | 2/32 | 0.001595 | 0.025421 |
| BP | negative regulation of cardiac muscle hypertrophy | 2/32 | 0.001687 | 0.026207 |
| BP | negative regulation of GTPase activity | 2/32 | 0.001687 | 0.026207 |
| BP | cellular response to lipoprotein particle stimulus | 2/32 | 0.001782 | 0.02733 |
| BP | negative regulation of muscle hypertrophy | 2/32 | 0.001879 | 0.028462 |
| BP | positive regulation of sterol transport | 2/32 | 0.001978 | 0.029062 |
| BP | positive regulation of cholesterol transport | 2/32 | 0.001978 | 0.029062 |
| BP | negative regulation of angiogenesis | 3/32 | 0.002013 | 0.029062 |
| BP | regulation of reactive oxygen species metabolic process | 3/32 | 0.002013 | 0.029062 |
| BP | negative regulation of blood vessel morphogenesis | 3/32 | 0.002091 | 0.029836 |
| BP | negative regulation of vasculature development | 3/32 | 0.002131 | 0.030053 |
| BP | positive regulation of inflammatory response | 3/32 | 0.002212 | 0.03049 |
| BP | detoxification | 3/32 | 0.002212 | 0.03049 |
| BP | positive regulation of smooth muscle cell migration | 2/32 | 0.002512 | 0.034252 |
| BP | regulation of angiogenesis | 4/32 | 0.00273 | 0.03619 |
| BP | negative regulation of lymphocyte activation | 3/32 | 0.002736 | 0.03619 |
| BP | regulation of activated T cell proliferation | 2/32 | 0.002743 | 0.03619 |
| BP | regulation of vasculature development | 4/32 | 0.002903 | 0.037893 |
| BP | acute-phase response | 2/32 | 0.002983 | 0.03853 |
| BP | activated T cell proliferation | 2/32 | 0.003107 | 0.039125 |
| BP | negative regulation of reactive oxygen species metabolic process | 2/32 | 0.003107 | 0.039125 |
| BP | positive regulation of endopeptidase activity | 3/32 | 0.003125 | 0.039125 |
| BP | vascular endothelial cell proliferation | 2/32 | 0.003361 | 0.041243 |
| BP | regulation of vascular endothelial cell proliferation | 2/32 | 0.003361 | 0.041243 |
| BP | cellular defense response | 2/32 | 0.003492 | 0.041963 |
| BP | adaptive immune response based on somatic recombination of immune receptors built from immunoglobulin superfamily domains | 4/32 | 0.003533 | 0.041963 |
| BP | regulation of T cell proliferation | 3/32 | 0.003546 | 0.041963 |
| BP | regulation of T cell activation | 4/32 | 0.003567 | 0.041963 |
| BP | receptor signaling pathway via STAT | 3/32 | 0.003601 | 0.041963 |
| BP | regulation of cholesterol efflux | 2/32 | 0.003625 | 0.041963 |
| BP | peptidyl-tyrosine phosphorylation | 4/32 | 0.003704 | 0.04248 |
| BP | regulation of lipid storage | 2/32 | 0.003761 | 0.042488 |
| BP | peptidyl-tyrosine modification | 4/32 | 0.003774 | 0.042488 |
| BP | cellular response to cAMP | 2/32 | 0.003898 | 0.043485 |
| BP | positive regulation of peptidase activity | 3/32 | 0.004241 | 0.046877 |
| BP | positive regulation of protein tyrosine kinase activity | 2/32 | 0.004473 | 0.048747 |
| BP | negative regulation of leukocyte activation | 3/32 | 0.004489 | 0.048747 |
| BP | response to reactive oxygen species | 3/32 | 0.004879 | 0.052509 |
| BP | leukocyte cell-cell adhesion | 4/32 | 0.00502 | 0.053562 |
| BP | positive regulation of protein kinase activity | 4/32 | 0.005281 | 0.05586 |
| BP | negative regulation of lipid localization | 2/32 | 0.005403 | 0.056447 |
| BP | T cell proliferation | 3/32 | 0.005429 | 0.056447 |
| BP | regulation of endopeptidase activity | 4/32 | 0.005643 | 0.058184 |
| BP | cellular ketone metabolic process | 3/32 | 0.005865 | 0.05947 |
| BP | protein kinase B signaling | 3/32 | 0.005865 | 0.05947 |
| BP | regulation of cardiac muscle hypertrophy | 2/32 | 0.006069 | 0.060754 |
| BP | negative regulation of cell activation | 3/32 | 0.00609 | 0.060754 |
| BP | cholesterol efflux | 2/32 | 0.006415 | 0.063474 |
| BP | regulation of muscle hypertrophy | 2/32 | 0.006591 | 0.063679 |
| BP | positive regulation of fat cell differentiation | 2/32 | 0.006591 | 0.063679 |
| BP | regulation of cellular response to insulin stimulus | 2/32 | 0.006591 | 0.063679 |
| BP | regulation of DNA-binding transcription factor activity | 4/32 | 0.006934 | 0.066109 |
| BP | positive regulation of phagocytosis | 2/32 | 0.00695 | 0.066109 |
| BP | regulation of peptidase activity | 4/32 | 0.007203 | 0.067515 |
| BP | reactive oxygen species metabolic process | 3/32 | 0.007212 | 0.067515 |
| BP | negative regulation of endothelial cell proliferation | 2/32 | 0.007318 | 0.067515 |
| BP | nitric oxide biosynthetic process | 2/32 | 0.007318 | 0.067515 |
| BP | activation of cysteine-type endopeptidase activity involved in apoptotic process | 2/32 | 0.007506 | 0.068658 |
| BP | regulation of lymphocyte proliferation | 3/32 | 0.007554 | 0.068658 |
| BP | regulation of mononuclear cell proliferation | 3/32 | 0.007905 | 0.071321 |
| BP | mononuclear cell differentiation | 4/32 | 0.007993 | 0.071583 |
| BP | regulation of sterol transport | 2/32 | 0.008276 | 0.073056 |
| BP | regulation of cholesterol transport | 2/32 | 0.008276 | 0.073056 |
| BP | nitric oxide metabolic process | 2/32 | 0.008474 | 0.073743 |
| BP | positive regulation of muscle cell differentiation | 2/32 | 0.008474 | 0.073743 |
| BP | reactive nitrogen species metabolic process | 2/32 | 0.008674 | 0.074953 |
| CC | external side of plasma membrane | 9/31 | 2.35E-08 | 1.20E-06 |
| CC | endocytic vesicle | 5/31 | 0.000175 | 0.00446 |
| CC | endocytic vesicle membrane | 3/31 | 0.003369 | 0.057268 |
| MF | carbohydrate binding | 6/29 | 3.72E-06 | 0.000506 |
| MF | immune receptor activity | 4/29 | 8.11E-05 | 0.005514 |
| MF | endonuclease activity, active with either ribo- or deoxyribonucleic acids and producing 3'-phosphomonoesters | 2/29 | 0.000248 | 0.007759 |
| MF | long-chain fatty acid binding | 2/29 | 0.000248 | 0.007759 |
| MF | cargo receptor activity | 3/29 | 0.000285 | 0.007759 |
| MF | MHC protein complex binding | 2/29 | 0.001457 | 0.033018 |
| MF | MHC protein binding | 2/29 | 0.001979 | 0.038453 |
| MF | scavenger receptor activity | 2/29 | 0.002473 | 0.040572 |
| MF | fatty acid binding | 2/29 | 0.002685 | 0.040572 |
| MF | lyase activity | 3/29 | 0.003683 | 0.050094 |
| MF | protein kinase regulator activity | 3/29 | 0.004753 | 0.058762 |
| MF | endoribonuclease activity | 2/29 | 0.005707 | 0.064674 |
| MF | kinase regulator activity | 3/29 | 0.006642 | 0.069484 |
| MF | monocarboxylic acid binding | 2/29 | 0.007171 | 0.069658 |
| MF | antioxidant activity | 2/29 | 0.00787 | 0.071359 |

* MF:molecular function; BP: biological process; CC: cellular components

Table S4. DO enrichment of 57 terms.

| Description | GeneRatio | BgRatio | pvalue | p.adjust |
| --- | --- | --- | --- | --- |
| Atherosclerosis | 8/24 | 364/10312 | 1.00E-06 | 0.00019 |
| Arteriosclerotic Cardiovascular Disease | 8/24 | 365/10312 | 1.02E-06 | 0.00019 |
| Arteriosclerosis | 8/24 | 411/10312 | 2.49E-06 | 0.00031 |
| Tuberculosis | 6/24 | 201/10312 | 5.11E-06 | 0.00048 |
| Intestinal Disease | 7/24 | 417/10312 | 3.20E-05 | 0.00242 |
| Primary Bacterial Infectious Disease | 6/24 | 295/10312 | 4.54E-05 | 0.00286 |
| Myocardial Infarction | 7/24 | 453/10312 | 5.44E-05 | 0.00294 |
| Bacterial Infectious Disease | 6/24 | 359/10312 | 1.35E-04 | 0.00638 |
| Lymphatic System Disease | 4/24 | 121/10312 | 1.60E-04 | 0.00671 |
| Malaria | 4/24 | 125/10312 | 1.81E-04 | 0.00685 |
| Hypersensitivity Reaction Type Iv Disease | 4/24 | 130/10312 | 2.11E-04 | 0.00724 |
| Hypersensitivity Reaction Disease | 4/24 | 152/10312 | 3.83E-04 | 0.01207 |
| Lymphadenitis | 3/24 | 70/10312 | 5.47E-04 | 0.01478 |
| Lymph Node Disease | 3/24 | 70/10312 | 5.47E-04 | 0.01478 |
| Pre-Eclampsia | 5/24 | 309/10312 | 6.22E-04 | 0.01567 |
| Parasitic Protozoa Infectious Disease | 4/24 | 178/10312 | 6.96E-04 | 0.01645 |
| Atopic Dermatitis | 4/24 | 183/10312 | 7.72E-04 | 0.01717 |
| Allergic Contact Dermatitis | 4/24 | 192/10312 | 9.24E-04 | 0.01941 |
| Contact Dermatitis | 4/24 | 195/10312 | 9.79E-04 | 0.01948 |
| Kidney Failure | 5/24 | 360/10312 | 1.24E-03 | 0.02241 |
| Parasitic Infectious Disease | 4/24 | 208/10312 | 1.24E-03 | 0.02241 |
| Esophageal Carcinoma | 4/24 | 219/10312 | 1.51E-03 | 0.02587 |
| Acute Myocardial Infarction | 3/24 | 105/10312 | 1.78E-03 | 0.02859 |
| Fatty Liver Disease | 4/24 | 232/10312 | 1.86E-03 | 0.02859 |
| Hyperandrogenism | 2/24 | 28/10312 | 1.89E-03 | 0.02859 |
| Acute Kidney Failure | 3/24 | 110/10312 | 2.03E-03 | 0.02952 |
| Sarcoidosis | 3/24 | 116/10312 | 2.36E-03 | 0.03104 |
| Dermatitis | 4/24 | 248/10312 | 2.37E-03 | 0.03104 |
| Allergic Rhinitis | 3/24 | 117/10312 | 2.42E-03 | 0.03104 |
| Female Reproductive System Disease | 5/24 | 421/10312 | 2.47E-03 | 0.03104 |
| Non-Alcoholic Fatty Liver Disease | 3/24 | 120/10312 | 0.002602 | 0.03104 |
| Esophageal Cancer | 4/24 | 255/10312 | 0.002627 | 0.03104 |
| Hepatitis C | 4/24 | 258/10312 | 0.002741 | 0.0314 |
| Hepatitis | 5/24 | 452/10312 | 0.003368 | 0.03744 |
| In Situ Carcinoma | 3/24 | 140/10312 | 0.004023 | 0.04345 |
| Lipid Storage Disease | 4/24 | 305/10312 | 0.004994 | 0.05244 |
| Rhinitis | 3/24 | 156/10312 | 0.005444 | 0.05397 |
| Nasal Cavity Disease | 3/24 | 157/10312 | 0.005541 | 0.05397 |
| Morbid Obesity | 2/24 | 49/10312 | 0.005711 | 0.05397 |
| Colonic Disease | 2/24 | 49/10312 | 0.005711 | 0.05397 |
| Nose Disease | 3/24 | 168/10312 | 0.006685 | 0.06066 |
| Pre-Malignant Neoplasm | 3/24 | 169/10312 | 0.006796 | 0.06066 |
| Esophagus Adenocarcinoma | 2/24 | 54/10312 | 0.0069 | 0.06066 |
| Cerebrovascular Disease | 4/24 | 337/10312 | 0.007094 | 0.06095 |
| Extrinsic Cardiomyopathy | 2/24 | 56/10312 | 0.007405 | 0.0622 |
| Lysosomal Storage Disease | 4/24 | 346/10312 | 0.007777 | 0.06391 |
| Liver Cirrhosis | 4/24 | 354/10312 | 0.008419 | 0.06695 |
| Congestive Heart Failure | 4/24 | 355/10312 | 0.008502 | 0.06695 |
| Alopecia | 2/24 | 62/10312 | 0.009016 | 0.06884 |
| Polycystic Ovary Syndrome | 3/24 | 188/10312 | 0.009105 | 0.06884 |
| Kawasaki Disease | 2/24 | 66/10312 | 0.010169 | 0.07273 |
| Pulmonary Fibrosis | 3/24 | 198/10312 | 0.010485 | 0.07273 |
| Bile Duct Adenocarcinoma | 3/24 | 198/10312 | 0.010485 | 0.07273 |
| Cholangiocarcinoma | 3/24 | 198/10312 | 0.010485 | 0.07273 |
| Ovarian Dysfunction | 3/24 | 199/10312 | 0.010629 | 0.07273 |
| Cerebral Infarction | 3/24 | 200/10312 | 0.010775 | 0.07273 |
| Upper Respiratory Tract Disease | 3/24 | 202/10312 | 0.011069 | 0.07341 |

Table S5. Primary weight of DEGs in the six classifications MLs.

| Genes | LASSO | RF | NN | GBM | DT | SVM |
| --- | --- | --- | --- | --- | --- | --- |
| ASGR2 | 1.25 | 5.9 | 1.99 | 247.23 | 6.41 | 5.07 |
| SOCS3 | 0.74 | 5.78 | -0.68 | 131.81 | 7.2 | 5.14 |
| AQP9 | 0 | 3.59 | 0.19 | 172.31 | 30.6 | 4.32 |
| PPARG | 0 | 4.48 | -0.3 | 254.65 | 7.63 | 4.21 |
| RNASE1 | 0.51 | 4.36 | -0.44 | 62.1 | 2.94 | 6.18 |
| DYSF | 0 | 1.17 | 1.35 | 3.42 | 17.48 | 6.04 |
| S100A9 | 0 | 3.11 | 0.18 | 2.45 | 22.6 | 5.24 |
| FCGR1A | 0 | 1.02 | 1.01 | 0.92 | 17.48 | 5.71 |
| VSIG4 | 0.01 | 2.59 | 0.6 | 25 | 5.87 | 7.2 |
| STAB1 | 0 | 2.78 | 1.16 | 11.78 | 4.34 | 5.08 |
| MYBL1 | -0.33 | 4.04 | 0.3 | 49.87 | 0 | 4.37 |
| KLRD1 | 0 | 1.52 | -1.3 | 2.81 | 1.44 | 6.13 |
| ECRP | 0.42 | 2.59 | -0.47 | 29.13 | 0 | 4.56 |
| TCN2 | 0 | 2.7 | 0.53 | 17.88 | 0 | 6.51 |
| FAM20A | 0 | 1.85 | -0.15 | 37.53 | 0 | 8.37 |
| MERTK | 0.17 | 1.14 | -0.42 | 3.37 | 3.09 | 5.97 |
| HP | 0 | 0.51 | -1.56 | 0.78 | 0 | 3.77 |
| RNASE2 | 0 | 0.97 | 0.84 | 3.3 | 0 | 5.82 |
| DTHD1 | 0 | 0.75 | -0.9 | 12.35 | 0 | 5.51 |
| CLC | 0 | 0.63 | 1.43 | 5.68 | 0 | 3.02 |
| SNORD20 | -0.12 | 0.82 | -0.48 | 1.68 | 3.91 | 4.19 |
| CD163 | 0 | 0.91 | 0.57 | 0.16 | 3.42 | 4.73 |
| NRG1 | 0 | 1.2 | 0.49 | 4.42 | 0 | 5.3 |
| SNORD45B | 0 | 0.72 | 1.27 | 3.18 | 0 | 2.79 |
| CYP1B1 | 0 | 0.84 | 0.5 | 0.31 | 0 | 5.5 |
| KLRC2 | 0 | 0.44 | -1.02 | 0.4 | 0 | 3.83 |
| TMEM176A | 0 | 0.48 | 1.33 | 0.4 | 0 | 2.03 |
| SLED1 | 0 | 0.51 | 0.48 | 4.55 | 1.44 | 4.1 |
| FCGR1A.2 | 0 | 1.38 | 0 | 0.58 | 18.94 | 0 |
| SERPINB2 | 0 | 0.49 | 0.42 | 0.66 | 0 | 4.52 |
| FCGR1A.1 | 0 | 1.09 | 0 | 0.25 | 18.94 | 0 |
| KLRC4 | 0 | 0.75 | -0.42 | 0.06 | 0 | 3.59 |
| KLRA1P | 0 | 0.58 | -0.13 | 0.05 | 2.32 | 4.29 |
| MIR21 | 0 | 0.46 | -0.18 | 2.45 | 0 | 4.18 |
| CES1 | 0 | 0.71 | -0.1 | 8.64 | 0 | 3.92 |
| KLRC4-KLRK1 | 0 | 0.41 | 0 | 0.68 | 2.32 | 3.64 |
| KLRC3 | 0 | 0.43 | -0.19 | 0.18 | 0 | 3.29 |
| NRG1.1 | 0 | 0.78 | 0 | 0.62 | 0 | 0 |
| FMN1.1 | 0 | 0.42 | 0 | 1.42 | 0 | 0 |

*LASSO, Least Absolute Shrinkage and Selection Operator; RF, Random Forest; GBM, Gradient Boosting Machine; DT, Decision Trees ; NN, Neural Network.

Fig.S1 The difference between the 22 immune cells.

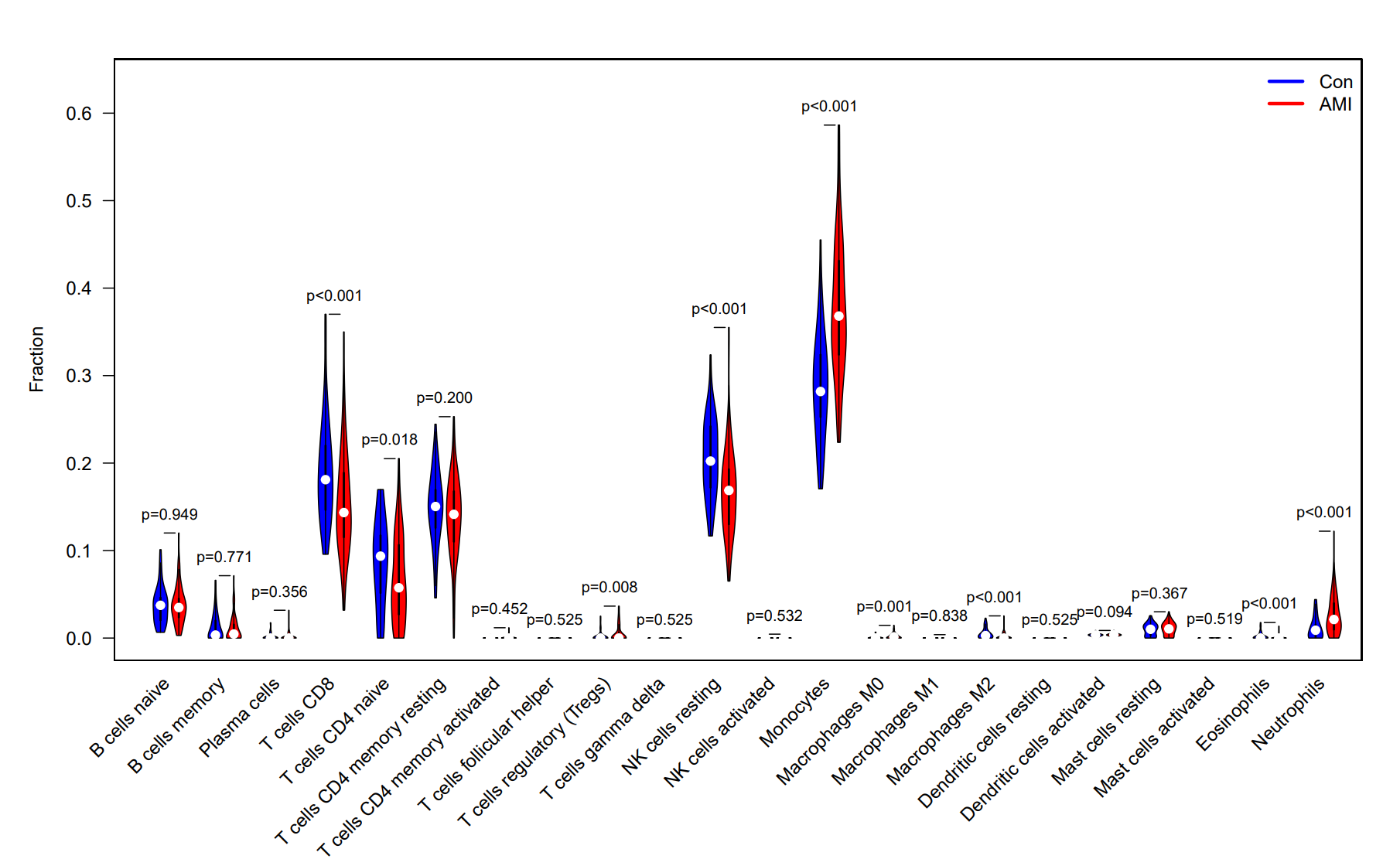

Fig S2. The other 7 immune cells correlation analysis of SOCS3.

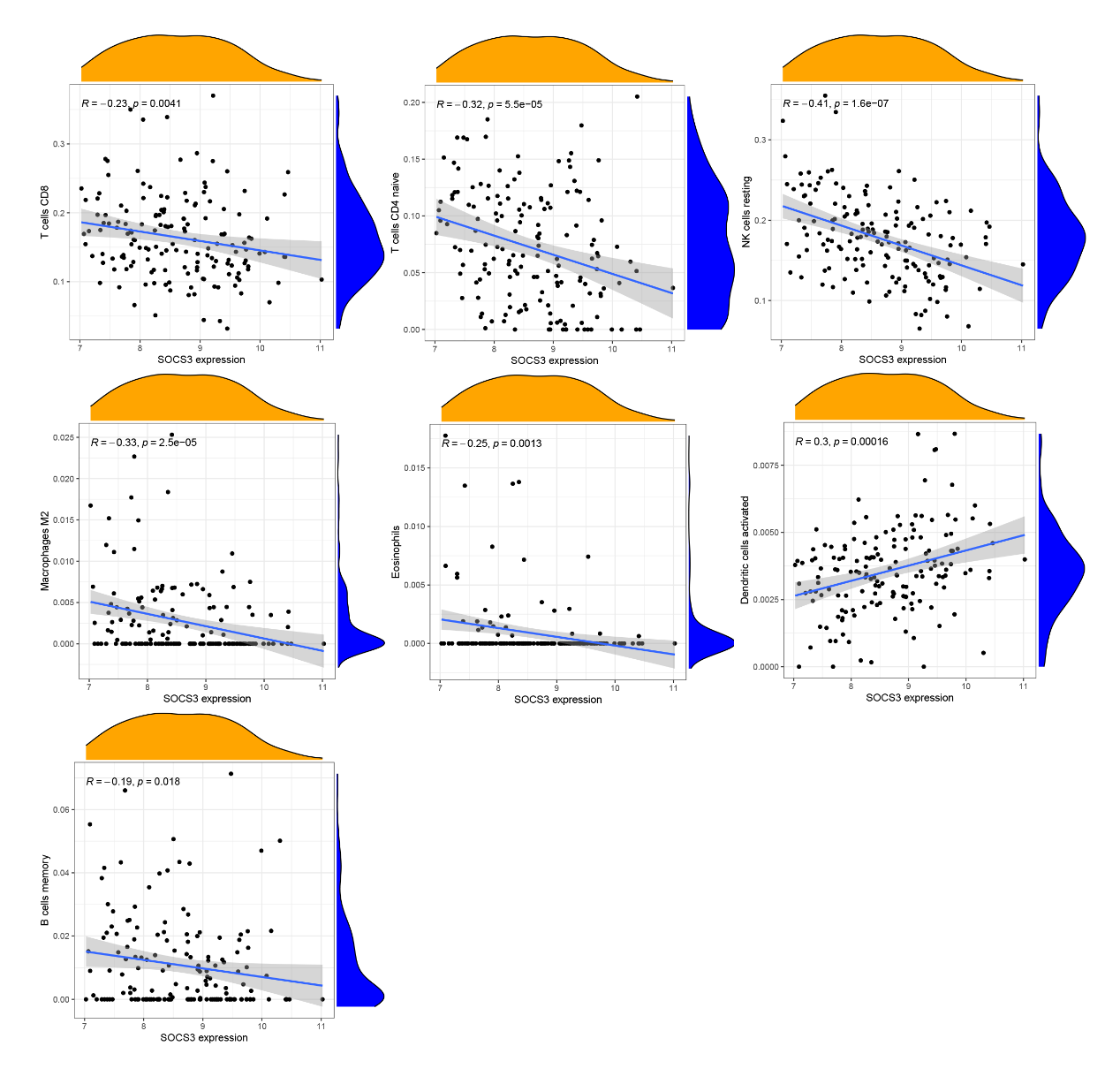

Fig S3. The other 7 immune cells correlation analysis of AQP9.

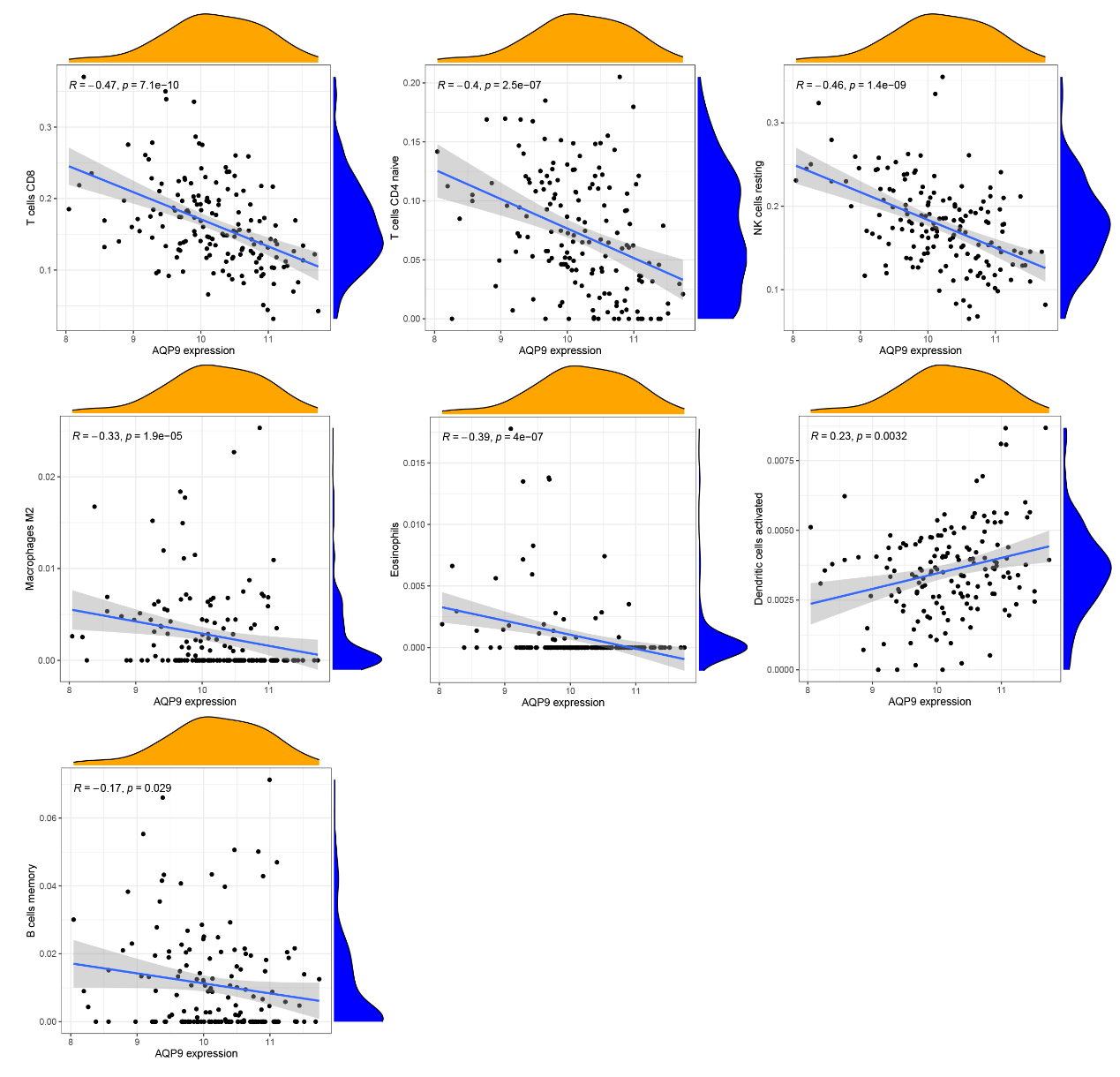

Fig S4. The Correlation analysis of 9 clinical variables.

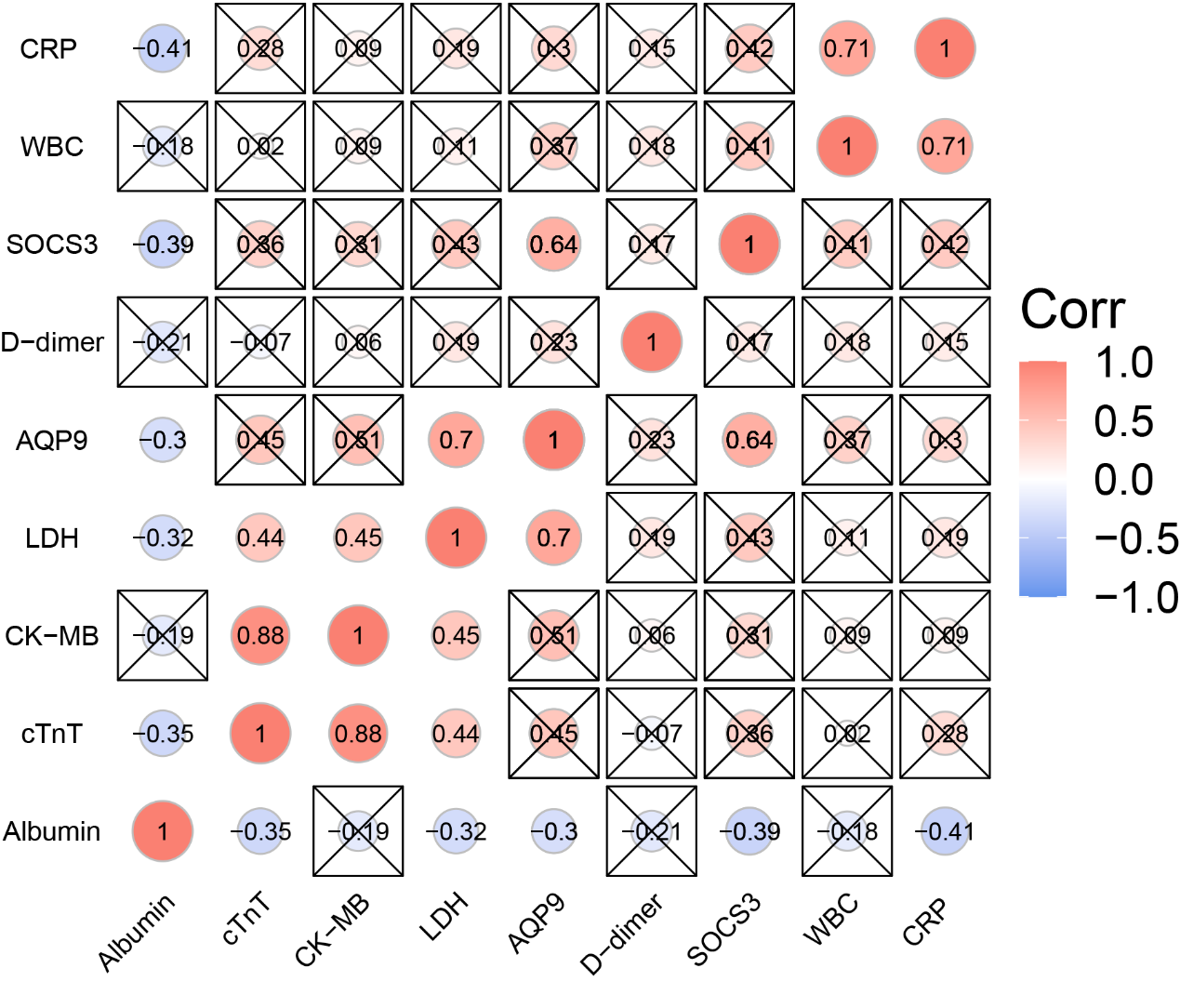
